# Endophenotype Effect Sizes Provide Evidence Supporting Variant Pathogenicity in Monogenic Disease Susceptibility Genes

**DOI:** 10.1101/2021.08.28.21262571

**Authors:** Valerie N. Morrill, Jennifer L. Halford, Seung Hoan Choi, Sean J. Jurgens, Giorgio Melloni, Nicholas A. Marston, Lu-Chen Weng, Victor Nauffal, Amelia W. Hall, Sophia Gunn, Christina A. Austin-Tse, James P. Pirruccello, Shaan Khurshid, Heidi L. Rehm, Emelia J. Benjamin, Eric Boerwinkle, Jennifer A. Brody, Adolfo Correa, Brandon K. Fornwalt, Namrata Gupta, Christopher M. Haggerty, Stephanie Harris, Susan R. Heckbert, Charles C. Hong, Charles Kooperberg, Henry J. Lin, Ruth J. F. Loos, Braxton D. Mitchell, Alanna C. Morrison, Wendy Post, Bruce M. Psaty, Susan Redline, Kenneth M Rice, Stephen S. Rich, Jerome I. Rotter, Peter F. Schnatz, Elsayed Z. Soliman, Nona Sotoodehnia, Eugene K. Wong, NHLBI Trans-Omics for Precision Medicine (TOPMed) Consortium, Marc S. Sabatine, Christian T. Ruff, Kathryn L. Lunetta, Patrick T. Ellinor, Steven A. Lubitz

## Abstract

Accurate and efficient classification of variant pathogenicity is critical for research and clinical care. Using data from three large studies, we demonstrate that population-based associations between rare variants and quantitative endophenotypes for three monogenic diseases (low-density-lipoprotein cholesterol for familial hypercholesterolemia, electrocardiographic QTc interval for long QT syndrome, and glycosylated hemoglobin for maturity-onset diabetes of the young) are proxies for variant pathogenicity. Effect sizes were associated with pathogenic ClinVar assertions (*P*<0.001 for each trait) and discriminated pathogenic from non-pathogenic variants (area under the curve 0.82-0.83 across endophenotypes). Large effect size thresholds provided evidence supporting pathogenicity for up to 35% of rare variants of uncertain significance or not in ClinVar in disease susceptibility genes, using an effect size threshold of ≥ 0.5 times the endophenotype standard deviation. We propose that variant associations with quantitative endophenotypes for monogenic diseases can provide evidence supporting pathogenicity.

---

Determining the clinical significance of rare genetic variation has critical implications for research and optimal care for patients and their families.^1^ Incorrect classification of genetic variation, though rare, may place patients and their relatives at risk for adverse consequences of disease, inappropriate therapies with related complications, and anxiety.^2^ Insufficient evidence for pathogenicity is common and the inability to discriminate variants of uncertain significance (VUS) remains a significant barrier.^3^ In clinical practice, genetic testing is frequently hampered by discovery of VUS and conflicting variant classifications between laboratories.^4–6^ Although a process of variant classification advanced by the American College of Medical Genetics (ACMG) and Association for Molecular Pathology (AMP) exists,^7^ it is laborious and prone to adjudicator disagreement.^8,9^ For example, one study across three laboratories found low concordance (Cohen K=0.26) in classifying variants in *SCN5A* and *KCNH2* – two disease-causing genes for the long QT syndrome (LQTS) which can cause sudden cardiac death.^10^ Nevertheless, with increasing use of genetic sequencing in both research and clinical practice there is now a rapidly growing pool of rare variants requiring adjudication.

Quantitative endophenotypes are inherently more powerful phenotypes for genetic association than dichotomous disease status indicators, and often may be more easily ascertained. The emergence of large-scale human genetic sequence and phenotype data in biorepositories provides an opportunity to assess whether endophenotypes for monogenic diseases can be leveraged to enable scalable and accurate variant pathogenicity assertions. Currently, variant classification practices do not account for rare variant associations with endophenotypes for monogenic diseases. We hypothesized that quantitative endophenotypes for monogenic diseases measured at population-scale can provide evidence of variant pathogenicity.

To assess our hypothesis, we studied three monogenic diseases for which easily ascertained human-derived endophenotypes exist, and ambiguous variant classification is an established challenge.^2,4,6,11^ Familial hypercholesterolemia (FH) is the most common inherited disorder in medicine, affecting about 1 in 250 individuals. FH is characterized by elevated blood levels of low-density lipoprotein cholesterol (LDL-C), which may lead to premature coronary artery disease and myocardial infarction.^3^ LQTS is a common cause of arrhythmias that affects about 1 in 2000 individuals and may lead to sudden cardiac death.^12^ The electrocardiographic corrected QT interval (QTc) is an easily measured endophenotype for LQTS. Maturity-onset diabetes of the young (MODY) is a monogenic form of diabetes often misdiagnosed as type 1 or type 2 diabetes that affects about 1 in 10,000 adults.^13^ Glycosylated hemoglobin (HbA1c) levels are a routinely measured blood biomarker for diabetes.

We studied the relations between rare coding variants, defined here as those with a minor allele frequency (MAF) <1%, in disease-causing genes for FH and MODY with LDL-C and HbA1c levels, respectively, among individuals who underwent whole exome sequencing (WES) in the UK Biobank (UKBB) and replicated findings in the Further Cardiovascular Outcomes Research With PCSK9 Inhibition in Subjects With Elevated Risk (FOURIER) randomized controlled trial.^14^ We studied the relations between rare variants in LQTS susceptibility genes and QTc intervals among individuals who underwent WES in the UKBB and replicated findings in the National Heart Lung and Blood Institute’s (NHLBI) Trans-Omics for Precision Medicine (TOPMed) program,^15^ in which samples underwent whole genome sequencing (WGS).

## RESULTS

### Sample characteristics

Our discovery analysis included multi-ancestry samples from the UKBB comprising 189,656 participants with LDL-C measurements, 33,521 with QTc measurements, and 189,744 with HbA1c measurements (**Supplemental Figures 1-3**). Sample characteristics are reported in **Table 1**. Our replication analyses for the LDL-C and HbA1c endophenotypes included 14,038 and 12,798 samples, respectively, from the FOURIER trial. Replication analyses for the QTc endophenotype included 26,976 individuals from TOPMed (Replication sample characteristics are reported in **Supplemental Table 1**). A sensitivity analysis including individuals in the UKBB of European ancestry included 165,786 participants with LDL-C measurements, 28,249 with QTc measurements, and 166,377 with HbA1c measurements (European sample characteristics are reported in **Supplemental Table 2**). A schema of the study design is shown in **Figure 1**.

**Table 1.** Baseline cohort characteristics by endophenotype in the UK Biobank

| **Characteristic** | **LDL-C (mg/dL)** | **QTc (ms)** | **HbA1c (%)** |
| --- | --- | --- | --- |
| Participants with a measurable endophenotype, n | 189,656 | 33,521 | 189,744 |
| Median endophenotype value (Q1-Q3) | 136 (114 - 159) | 411 (396 - 426) | 5.4 (5.1 – 5.6) |
| Male, n (%) | 85,200 (44.9) | 15,583 (49.4) | 85,212 (44.9) |
| European ancestry, n (%) | 165,786 (87.4) | 28,249 (84.3) | 166,337 (87.7) |
| Mean age, years (SD) | 56.9 (8.1) | 52.5 (5.7) | 57.0 (8.1) |
| Myocardial infarction, n (%) | 2,995 (1.5) | 483 (1.4) | - |
| Statin usage, n (%) | 31,910 (16.8) | - | - |
| Mean high-density lipoprotein, mg/dL (SD) | 56.3 (14.2) | - | - |
| Heart failure, n (%) | - | 74 (0.2) | - |
| Beta blocker usage, n (%) | - | 1,701 (5.1) | - |
| Calcium channel blocker usage, n (%) | - | 2,455 (7.3) | - |
| Type 2 diabetes medication usage, n (%) | - | - | 7,090 (3.7) |
| Mean corpuscular volume, femtoliters (SD) | - | - | 91.2 (4.5) |
| NB: only select relevant characteristics for the given monogenic disease of interest are displayed |  |  |  |

**Figure 1.**
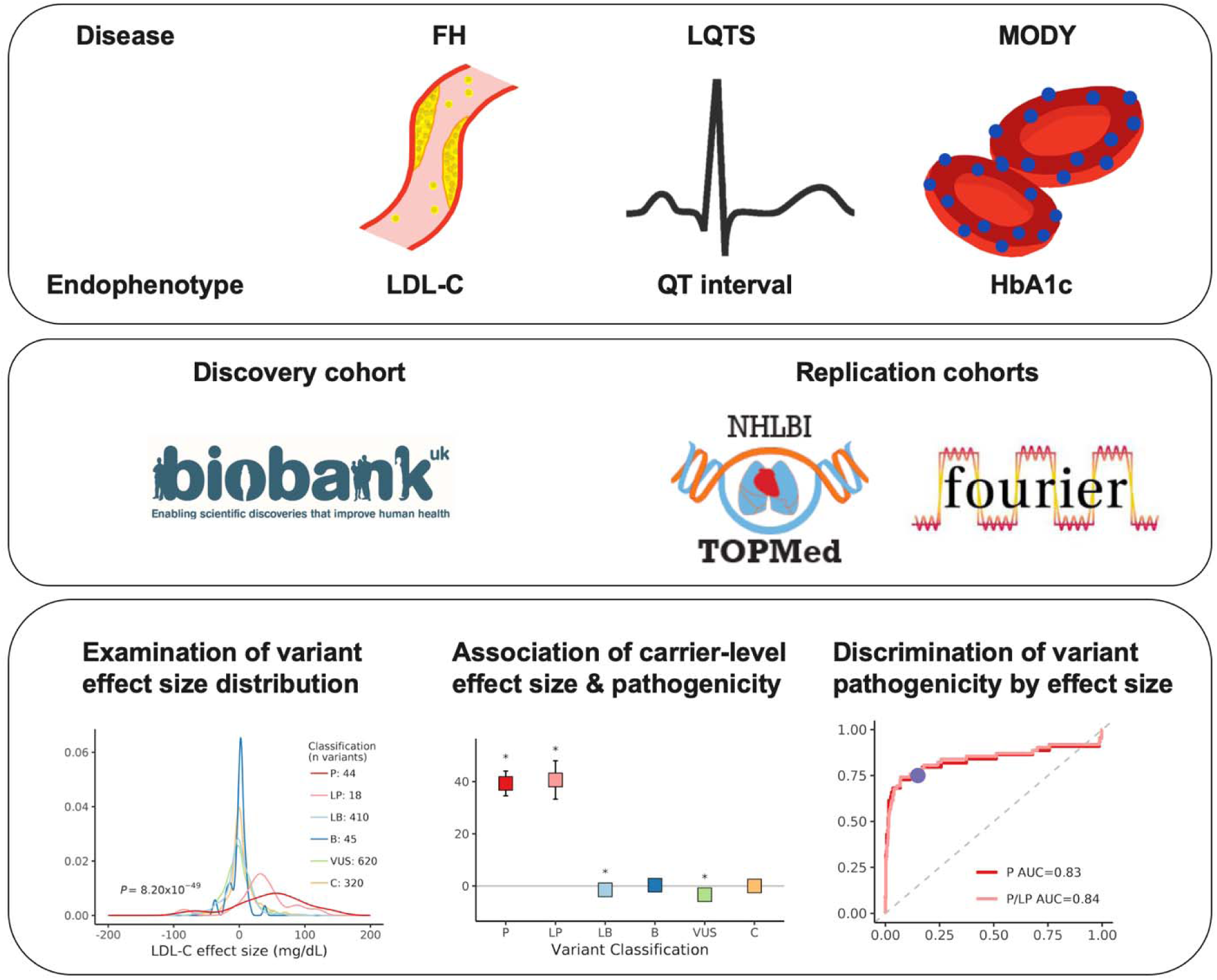
Schema of study design.

### Variant-level effect sizes and pathogenicity category

Within definitive FH genes (*LDLR, APOB, PCSK9*)^16^, we observed 3,487 rare (within-sample MAF <1%) coding variants in the UKBB; of these, 1,457 (41.7%) were present in ClinVar with clinical pathogenicity assertions. Single variant association testing between each variant in the FH genes and LDL-C levels produced variant effect sizes that differed by pathogenicity category. Pathogenic and likely pathogenic variants exhibited the largest mean effect sizes (effect size ± standard deviation [SD]: 47.02 ± 53.10 and 44.50 ± 58.43 milligrams per deciliter [mg/dL], respectively) (**Figure 2A**). For variants within definitive LQTS genes (*KCNQ1, KCNH2, SCN5A*)^15^, we observed 1,083 rare coding variants in the UKBB, 764 (70.5%) of which were present in ClinVar. Variant effect sizes for QTc duration differed across categories with pathogenic and likely pathogenic variants exhibiting the largest mean effect sizes (30.10 ± 29.90 and 11.54 ± 36.98 milliseconds [ms], respectively) (**Figure 2B**). For variants within the common MODY genes (*GCK, HNF1A, HNF1B, HNF4A*)^13^, we observed 1,046 rare coding variants in the UKBB; of these, 187 (17.9%) were present in ClinVar. Variant effect sizes for HbA1c differed across categories with pathogenic and likely pathogenic variants exhibiting the largest mean effect sizes (0.70 ± 0.70 and 0.45 ± 0.37 %, respectively) (**Figure 2C**).

**Figure 2.**
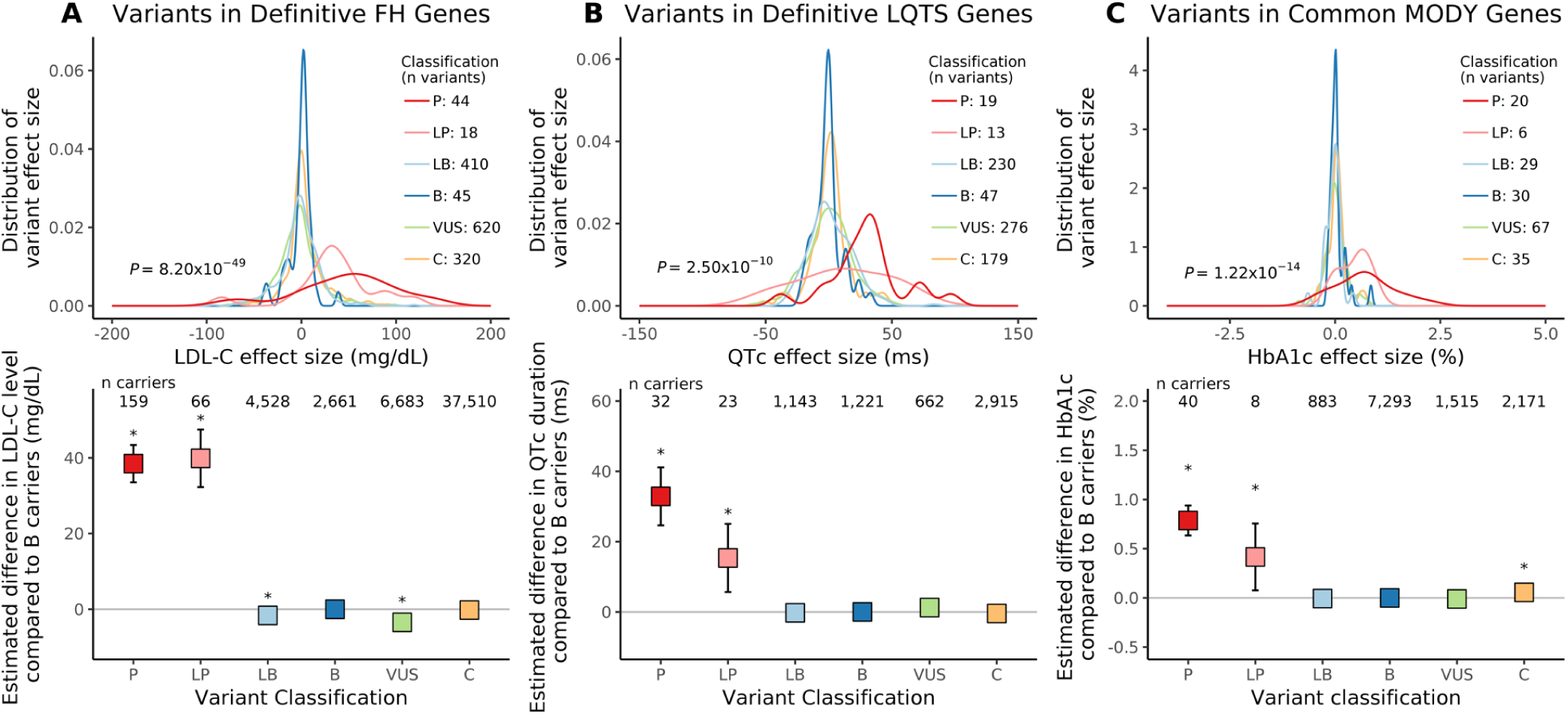
Association between effect size and variant pathogenicity for three monogenic disease endophenotypes. Panels A, B, and C display data for the LDL-C, QTc, and HbA1c endophenotypes, respectively, for rare variants found in the UK Biobank. Definitive familial hypercholesterolemia (FH) genes include *LDLR, APOB, PCSK9*; definitive long-QT syndrome (LQTS) genes include *KCNQ1, KCNH2, SCN5A*; common maturity-onset diabetes of the young (MODY) genes include *HNF1A, HNF1B, HNF4A, GCK*. Row 1 in each panel displays the variant effect size distribution by ClinVar pathogenicity category (colored). Row 2 in each panel displays the estimated difference in endophenotype value comparing carriers of a variant in each ClinVar pathogenicity category to carriers of benign variants (boxes) and 95% confidence intervals (whiskers); * indicates *P* < 0.05. Variant classification includes B: benign, LB: likely benign, LP: likely pathogenic, P: pathogenic, VUS: variant of uncertain significance, C: conflicting.

Results were similar in sensitivity analyses using expanded gene sets for FH, LQTS, and MODY informed by commercially available panels (**Supplemental Tables 3-5**)^17–19^. The distributions of effect sizes for variants tested for association with LDL-C, QTc, and HbA1c did not differ across pathogenicity categories in a control panel of hereditary cancer genes^20^, which would not be expected to be associated with either LDL-C levels, QTc duration, or HbA1c percentage (**Supplemental Figure 4**).

### Carrier-level effect sizes and pathogenicity category

We then assessed the estimated differences in endophenotype measurements for carriers of variants in each pathogenicity category compared to carriers of benign variants. Compared to carriers of benign variants, LDL-C levels were greater among carriers of pathogenic (difference 38.5 mg/dL, 95% CI 33.5, 43.4, *P* =1.08×10^−52^) and likely pathogenic (difference 39.9 mg/dL, 95% CI 32.3, 47.5, *P* = 8.44×10^−25^) variants in FH genes (**Figure 2A**). Similarly, QTc duration was greater among carriers of pathogenic (difference 32.9 ms, 95% CI 24.6, 41.1, *P* = 6.34×10^−15^) and likely pathogenic (difference 15.4 ms, 95% CI 5.7, 25.0, *P* = 1.89×10^−3^) variants compared to benign variant carriers (**Figure 2B**). Lastly, HbA1c percentage was greater among carriers of pathogenic (difference 0.79%, 95% CI 0.63, 0.94, *P* = 5.63×10^−24^) and likely pathogenic (difference 0.42%, 95% CI 0.08,0.76, *P* = 0.02) variants compared to benign variant carriers (**Figure 2C**). No significant differences in LDL-C, QTc, or HbA1c measures were observed among carriers of pathogenic or likely pathogenic variants in a control set of hereditary cancer genes (**Supplemental Figure 4**). In sensitivity analyses restricted to individuals of European ancestry in the UKBB (n=165,786 participants for LDL-C levels, n=28,249 participants for QTc duration, n=166,337 participants for HbA1c), we again observed that carriers of pathogenic and likely pathogenic variants had higher LDL-C levels, longer QTc intervals, and higher HbA1c percentages than carriers of benign variants (**Supplemental Figure 5**). A sensitivity analysis including only variants with minor allele counts ≤ 5 for association with the endophenotype demonstrated similar findings (**Supplemental Figure 6**).

### Replication of associations between effect sizes and pathogenicity category

We observed similar relations between rare variant effect sizes for each endophenotype and variant pathogenicity categories (**Supplemental Tables 6-8, Supplemental Figure 7**). Carriers of pathogenic variants had significantly greater endophenotype values compared to benign variant carriers for all endophenotypes.

### Variant-level effect sizes and discrimination of pathogenicity category

We then examined whether variant effect sizes can discriminate variant pathogenicity assertions. For the LDL-C endophenotype, variant effect sizes among FH genes discriminated pathogenic variants from variants classified as likely benign and benign in a logistic regression model with an AUC of 0.83 (95% CI 0.74, 0.92) and 0.91 (95% CI 0.87, 0.96) in the UKBB and FOURIER, respectively (**Figure 3A**). For the QTc interval, variant effect sizes among LQTS genes discriminated pathogenic variants with an AUC of 0.83 (95% CI 0.71, 0.96) and 0.79 (95% CI 0.66, 0.93) in the UKBB and TOPMed, respectively (**Figure 3B**). For HbA1c, variant effect sizes among MODY genes discriminated pathogenic variants with an AUC of 0.82 (95% CI 0.67, 0.97) and 0.92 (95% CI 0.79, 1) in the UKBB and FOURIER, respectively **(Figure 3C**). Slightly lower discrimination was observed when pathogenic and likely pathogenic variants were grouped together (**Figure 3**). We observed similar results in a sensitivity analysis in which we repeated the analysis after excluding overlapping variants from the replication cohorts that were present in the UKBB discovery cohort (**Supplemental Figure 8**). For analyses of LDL-C, 28.3% variants in definitive FH genes overlapped between the UKBB and FOURIER; for analyses of QTc, 62.0% of variants in definitive LQTS genes overlapped between the UKBB and TOPMed; for analyses of HbA1c, 45.0% of variants in common MODY genes overlapped between the UKBB and FOURIER.

**Figure 3.**
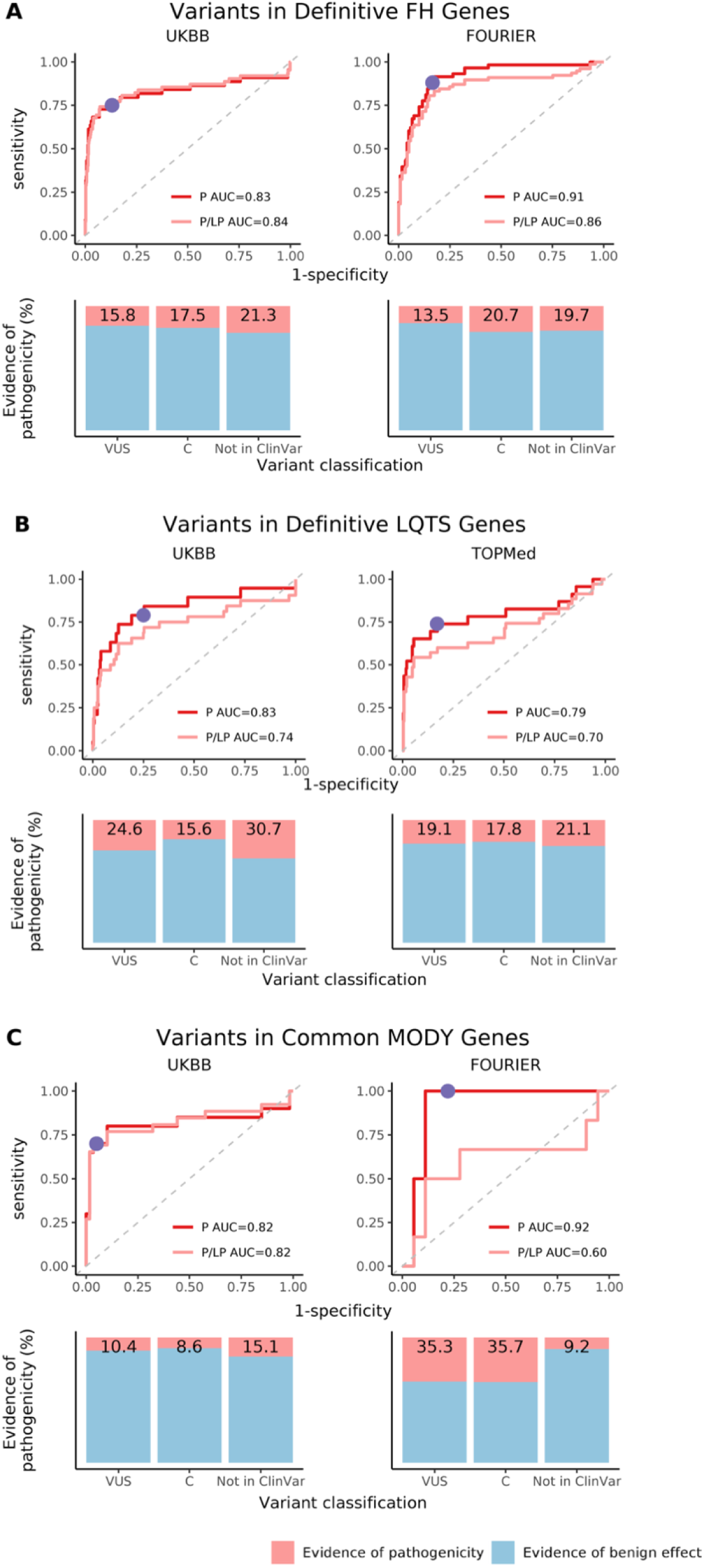
Discrimination of variant pathogenicity by effect size and percent of variants with evidence of pathogenicity. Panel A includes variants within familial hypercholesterolemia (FH) genes recommended for secondary findings return (*LDLR, APOB, PCSK9*); Panel B includes variants within the long-QT syndrome (LQTS) genes recommended for which the return of a secondary variant finding is endorsed (*KCNQ1, KCNH2, SCN5A)*; Panel C includes variants within common maturity-onset diabetes of the young (MODY) genes (*HNF1A, HNF1B, HNF4A, GCK*). Column 1 includes variants from the UK Biobank, the primary cohort; Column 2 includes variants from FOURIER and TOPMed, the replication cohorts. Rows 1, 3, and 5 display receiver operating characteristic (ROC) curves depicting discrimination of pathogenic variants according to effect size. Variant pathogenicity is taken from ClinVar. The red curve includes “Pathogenic” variants only; the pink curve includes “Pathogenic” and “Likely pathogenic” variants combined; area under the curve (AUC) values and variant numbers are reported in the legend. High effect size, defined as effect size greater than 0.5 standard deviations (SD) of the trait distribution, is plotted in purple on the “Pathogenic” variant ROC curve. Rows 2, 4, and 6 display variants by ClinVar category and shows the percent of variants with evidence for pathogenicity using the high effect size threshold defined in Rows 1, 3, and 5. Variants with evidence for pathogenicity, defined as having larger effect sizes than the threshold, are in pink; variants with evidence for a benign effect, defined as having smaller effect sizes than the threshold, are in light blue.

### Application of an effect size threshold to provide evidence of pathogenicity

Next, we selected a large effect size threshold which we applied to VUS or with conflicting assertions to document evidence for a pathogenic or benign classification. *A priori*, we selected variant effect size thresholds that corresponded to 0.5 SD of the endophenotype distribution in the UKBB. As a secondary analysis, we tabulated the sensitivity and specificity of other effect size thresholds based on a range of SD thresholds (**Supplemental Table 9**). For LDL-C levels, a large effect size threshold corresponding to 0.5 SD of the LDL-C distribution in the UKBB was 16.7 mg/dL. In the UKBB, the large LDL-C effect size threshold had a sensitivity of 75% and specificity of 87% for discriminating ClinVar designated pathogenic variants from non-pathogenic variants (likely benign, and benign). The large effect variant threshold in the UKBB provided evidence of pathogenicity for 15.8% of VUS and 17.5% of variants with conflicting assertions in FH genes (**Figure 3**). The large effect size threshold corresponding to 0.5 SD of the QTc distribution in the UKBB was 11.9 ms, which had a sensitivity of 79% and specificity of 75%. In the UKBB, this threshold provided evidence of pathogenicity for 24.6% of VUS and 15.6% of conflicting variants in the LQTS genes (**Figure 3**). For HbA1c, the large effect threshold was 0.31%, with a sensitivity of 70% and specificity of 95%; this threshold provided evidence of pathogenicity for 10.4% of VUS and 8.6% of conflicting variants in the MODY genes (**Figure 3**). In aggregate, of the 268 VUS across FH, LQTS, and MODY genes for which large effect size thresholds provided evidence of pathogenicity, 255 (95%) were missense variants, 8 (3%) were synonymous, 3 (1%) were splice region variants, with the remainder a mix of different functional consequences. Detailed characteristics of these variants are listed in **Supplemental Table 10**.

We then applied the large variant effect size threshold to novel variants that were not previously submitted to ClinVar to nominate these variants as putatively pathogenic or benign. Of rare variants in FH genes, 58% were not previously reported in ClinVar. The large effect size threshold nominated 21.3% of these variants as putatively pathogenic in the UKBB (**Figure 3**). Similar results were observed in FOURIER. Of rare variants in LQTS genes, 29% were not reported in ClinVar. The large effect size threshold provided evidence for pathogenicity for 30.7% (**Figure 3**). Similar results were observed in TOPMed. For the rare variants in MODY, 82% were not previously reported in ClinVar, and the large effect size threshold provided evidence for pathogenicity for 15.1% of these variants (**Figure 3**). In total, large effect size thresholds provided evidence of pathogenicity for 843 variants not previously reported in ClinVar in FH, LQTS, and MODY genes. Of these, 556 (65.6%) were missense variants, 234 (27.8%) were synonymous variants, with the remainder a mix of different functional consequences. Additional details are provided in **Supplemental Table 11**.

## DISCUSSION

We observed that population-level associations between genetic variants and readily ascertainable endophenotypes for monogenic diseases are informative for classifying variant pathogenicity. Specifically, rare variant effect sizes derived from association testing with LDL-C levels, electrocardiographic QTc duration, and HbA1c levels discriminated pathogenic from non-pathogenic variants in susceptibility genes for FH, LQTS, and MODY, respectively. A large variant effect size threshold provided evidence for pathogenicity for up to 35% of variants previously classified as VUS or with conflicting classifications in ClinVar. Additionally, up to 30% of variants without ClinVar assertions had large effect sizes, providing evidence for pathogenicity for variants not previously subjected to rigorous clinical assertion processes.

Our findings have two main implications. First, quantitative endophenotypes for monogenic diseases that are measurable in large scale populations can be leveraged to infer rare variant pathogenicity. We demonstrated that effect sizes for rare variants in FH, LQTS, and MODY monogenic disease susceptibility genes with corresponding endophenotypes are proxies for variant pathogenicity in three large studies. Single-variant association testing in biobanks has become a widely used tool to explore the relations between rare variants and phenotypes of interest, yet it has primarily been used for genetic discovery purposes.^21,22^ Our findings suggest that referencing effect sizes from large-scale rare variant association testing may have applications beyond variant or gene discovery, aiding in variant classification. We anticipate that our findings will be widely applicable beyond FH, LQTS and MODY to additional quantitative endophenotypes for other monogenic diseases.

Second, variant effect sizes can be used as rapid and scalable discriminators of variant pathogenicity and may serve as tools to resolve variants with uncertain significance or conflicting classifications. Further, variant effect sizes may provide valuable evidence in the classification of novel variants. We anticipate that such applications may be utilized either prior to initiating a formal variant classification process or in conjunction with existing methods, which will require further prospective evaluation. The traditional variant classification process, which relies on familial segregation data and functional studies is time-consuming, labor-intensive, and subject to uncertainty.^7^ A growing landscape of computational tools are being deployed to predict pathogenicity through assessment of variant conservation and prediction of variant effects on protein function.^23,24^ However, current algorithms for classifying variant pathogenicity do not account for population-based genotype-phenotype associations, which can be interpreted as human-derived physiologic indicators of variant expressivity.

The large number of novel variants discovered with sequencing efforts highlights a need for rapid and scalable methods for assessing variant pathogenicity. Indeed, many individuals in our studies carried rare coding variants in disease susceptibility genes that have not previously been classified or submitted to ClinVar with clinical pathogenicity assertions (58.2% of rare FH variants, 29.6% of rare LQTS variants, and 82.1% of MODY variants in the UKBB). Further analyses are warranted to compare the predictive utility of allelic effect size for discriminating variant pathogenicity against widely-used computational tools, examine the prognostic implications of large-effect variation, test different effect size thresholds for determining potential pathogenicity, and discover easily ascertainable endophenotypes for other monogenic diseases to aid in variant classification.

To facilitate use of our results into potential clinical practice and research, we will make variant level association results from the present analysis publicly accessible in the Cardiovascular Disease Knowledge Portal.^25^ We expect that as the number of sequenced individuals grows, large compendia of variant effect sizes with endophenotypes may help classify variants as potentially pathogenic or benign, facilitating both research and clinical variant classification.

Our work must be considered in the context of the study design. The UKBB and FOURIER studies are predominantly of European ancestry and the results of our analysis may not be generalizable to all ancestries. However, the replication of associations in TOPMed, which comprises a more diverse ancestral distribution of participants suggest the results are robust. Greater ancestral diversity anticipated with future biobank efforts will further refine the ability to resolve relations between specific variants and pathogenicity. ClinVar includes variant submissions primarily from clinical laboratories in the United States^26–28^ and therefore existing variant pathogenicity assertions may not adequately represent non-European ancestral groups.

Increased availability and equity of genetic testing among diverse populations is needed. We used single time-point endophenotype measurements in our analyses; repeated measures may increase measurement precision and power, and warrant examination. Pathogenic variants were rare among the coding variants assessed and potentially subject to imprecise effect size estimation.^29^ Some of the genes in the genetic testing panels for FH and LQTS (*LDLRAP1, KCNQ1, KCNE1, TRDN*) are associated with both autosomal dominant and recessive disease; as such, calculated effect sizes from an additive genetic model may be biased toward the null.^30–32^ We acknowledge that variant pathogenicity is not a discrete entity, and that discrimination of variants as “pathogenic” or “benign” may belie probabilistic gradients of pathogenicity or penetrance. As such, we submit that use of effect sizes may enable more quantitative inferences of variant pathogenicity.

In conclusion, population-based genetic association testing for monogenic disease endophenotypes may enable scalable inferences that provide evidence supporting variant pathogenicity. Future analyses are warranted to test whether large effect size variants are associated with clinical outcomes, and whether variant effect size information can be implemented in variant pathogenicity assertion workflows.

## ONLINE METHODS

### Study participants

The UK Biobank (UKBB) is a large, national, prospective cohort of ∼500,000 individuals with detailed medical history, electronic health record, and genetic data.^33^ Participants were recruited from 22 centers across the UK between 2006-2010 and aged 40-69 years at recruitment. Our analysis focused on participants with whole-exome sequencing (WES) and QT intervals extracted from resting 3-lead ECGs prior to a bicycle exercise protocol (n = 33,521), participants with WES and LDL-C measured (n= 189,656), and participants with WES and HbA1c measured (n=189,744). Informed consent was obtained from all participants, and the UKBB received approval from the Research Ethics Committee (11/NW/0382). Our study was approved by the Mass General Brigham Human Research Committee and conducted using the UKBB Resource (Application 17488).

### Ascertainment of clinical measurements and covariates

Detailed WES protocols for the UKBB are available elsewhere.^34^ In brief, 20X whole exome sequencing was performed using IDT xGen Exome Research Panel v1.0 including supplemental probes, reads were aligned to human genome build GRCh38, joint genotype calling and initial variant QC was performed. Procedures for procuring data are discussed in detail on the UKBB website.^35–37^ Clinical exclusion criteria and covariates were ascertained using self-report, ICD-9 and ICD-10 codes, and operation codes (**Supplemental Methods**). The extracted QT intervals were corrected using the Bazett formula, defined as QTc = QT / √RR, for subsequent analyses.^38^ LDL-C levels and other disease-relevant biomarkers including HDL-C and MCV were collected at baseline from all UKBB participants and measured by enzymatic protective selection analysis on a Beckman Coulter AU5800 device. Hemoglobin A1c was measured from baseline blood tests via HPLC analysis on a Bio-Rad VARIANT II Turbo. Diabetes medication use was ascertained via self-report.

### Genotype, variant, and sample quality control

#### Genotype

The Genotype QC protocol for the UKBB is described elsewhere.^39^ In brief, genotypes were removed with total depth >200 or <10. Homozygous reference calls were removed if genotype quality was <20. Homozygous alternative calls were removed if the ratio of A1 depth + A2 depth and total depth was less than 0.9, the ratio of A2 depth and total depth was less than 0.9 or phred-scaled genotype likelihood was < 20. Heterozygous calls were removed if the ratio of A1 depth + A2 depth and total depth was less than 0.9, the ratio of A2 depth and total depth was less than 0.2 or phred-scaled genotype likelihood was <20.

#### Variant

UKBB variants were removed if they were in low complexity regions, had call rates <90%, failed the Hardy Weinberg Equilibrium test (*P* = < 1.0×10^−15^), or were monomorphic in the final dataset.

#### Sample

Duplicate individuals in the UKBB were identified with KING2 and removed if not a monozygotic twin. Genetically determined sex was calculated using high quality (MAF ≥ 0.1%, missingness ≤ 1%, Hardy Weinberg Equilibrium *P* = > 10^−6^) independent variants on the X chromosome. Samples were removed if their genetically determined sex did not match their reported sex. Samples were removed if they were outliers (outside of 8 standard deviations (SD) from the mean) for quality control metrics including heterozygosity homozygosity ratio, transition and transversion ratio, SNP and Indel ratio, and the number of singletons per sample. Genetically determined ancestry groups were defined using high-quality independent variants. We estimated five ancestry groups supervised by 1000G participants using ADMIXTURE version 1.3.0.^40^ We defined a sample as a member of the ancestry group if the probability of belonging to that ancestry group was greater than 80%. We defined a sample as a member of the “undetermined ancestry group” if the probability of belonging to any ancestry group was less than 80% (**Supplemental Methods**). We conducted our primary analysis in a multi-ancestry cohort, followed by a sensitivity analysis restricted to samples with genetically defined European ancestry.

### Clinical trait exclusions and covariates

For our LDL-C analysis, clinical covariates included high-density lipoprotein (HDL), history of myocardial infarction, and history of statin usage. We imputed participants with incomplete data for HDL to the median HDL value. We imputed participants with incomplete data for history of myocardial infarction and history of statin usage to no history. This resulted in a cohort of 189,656 participants in our multi-ancestry cohort in the UKBB (**Supplemental Figure 1**).

For our QTc analysis, participants with Wolff-Parkinson-White Syndrome (WPW), history of pacemaker placement, 2nd or 3rd degree atrioventricular (AV) block, history of class I or class III antiarrhythmic drug usage, and/or history of digoxin usage were excluded from the analysis.

ECGs with QRS duration >120 ms and/or heart rate <40 or >120 beats/minute were excluded. Clinical covariates included beta blocker use, calcium channel blocker use, history of myocardial infarction, and history of heart failure. We imputed participants with incomplete data for history of myocardial infarction and history of heart failure to no history. The above QC steps resulted in a cohort of 33,521 participants for our primary analysis in UKBB (**Supplemental Figure 2**).

For our HbA1c analysis, clinical covariates included mean corpuscular volume (MCV) and self-reported diabetes medication usage including common medications such as insulin, metformin, DPP-4 inhibitors, GLP-1 receptor agonists, SGLT-2 inhibitors, sulfonylureas, and thiazolidinediones. We imputed participants with incomplete data for MCV to the median MCV value. This resulted in a cohort of 189,744 participants in our multi-ancestry cohort in the UKBB (**Supplemental Figure 3**).

### Variant pathogenicity assertions reported in ClinVar

ClinVar is a public database of reported sequence variants and their relations with human phenotypes.^41^ We identified variants submitted to ClinVar from clinical genetic testing laboratories with the most recent pathogenicity assertion after 2015 and downloaded entries from https://ftp.ncbi.nlm.nih.gov/pub/clinvar/ on 11/28/2020. We utilized variant pathogenicity assertions that were submitted from the clinical genetic testing laboratories for further analysis. Pathogenicity categories included “benign”, “likely benign”, “likely pathogenic”, and “pathogenic.” For variants with an aggregate classification in ClinVar as “benign/likely benign” or “pathogenic/likely pathogenic”, the most recent submission classification was used to disaggregate the two classes. Variants in ClinVar classified as “conflicting” due to multiple assertions with conflicting classifications and “variants of uncertain significance” (VUS) were classified as such. Ultimately, our analysis included 6 clinical pathogenicity categories: pathogenic, likely pathogenic, likely benign, benign, VUS, and conflicting. We then merged the ClinVar dataset with those of the UKBB, FOURIER, and TOPMed datasets by aligning with GRCh38 positions.

### Statistical analyses

#### Single variant association testing

We first estimated the empirical kinship matrix and derived principal components of ancestry using high-quality (missingness < 10%, HWE > 0.001, MAF > 0.1) independent (pruned with a window size of 200 kb, step size of 100 kb, and r2 threshold of 0.05) variants. The kinship matrix was estimated using the “--make-rel” function in PLINK 2.0.^42^ Principal components of ancestry were estimated in an unrelated subset using PCAir.^43^ We then derived variant effect sizes by fitting linear mixed effects models for each variant compared to non-carriers of each variant in which we regressed the endophenotype on the variant dosage, adjusting for age, sex, clinical covariates, the first 12 principal components of ancestry, and fitting a variance component proportional to the empirical kinship matrix, as well as separate residual variances for each ancestral group. We used GENESIS version 2.14.3^44^ and R version 3.6^45^ and assumed an additive genetic model.

#### Relations between estimated variant effect size and pathogenicity

For our LDL-C analysis, we examined the relations between estimated variant effect sizes and clinical pathogenicity assertions for variants (1) within FH genes included in the ACMG list of genes recommended for secondary findings reporting (*LDLR, APOB, PCSK9*),^16^ (2) within a commercially available FH panel (*LDLR, APOB, PCSK9*, and *LDLRAP*)^17^ and (3) within a control panel comprising of genes from a commercially available hereditary cancer panel (**Supplemental Table 12**).^20^ For our QTc analysis, we examined the relationship between estimated variant effect sizes and clinical pathogenicity assertions for variants in established LQTS genes. We grouped variants as those residing (1) within a list of LQTS genes included in the ACMG list of genes recommended for secondary findings reporting (*KCNQ1, KCNH2, SCN5A*),^16,46^ (2) within a commercially available LQTS panel (*ANK2, CACNA1C, CALM1, CALM2, CALM3, CAV3, KCNE1, KCNE2, KCNH2, KCNJ2, KCNQ1, SCN5A, TRDN*),^18^ and (3) within a control panel comprising of genes from a commercially available hereditary cancer panel (**Supplemental Table 12**).^20^ For our HbA1c analysis, we examined the relationship between estimated variant effect sizes and clinical pathogenicity assertions for variants in established MODY genes. We grouped variants as those residing (1) within a list of established MODY genes (*GCK, HNF1A, HNF1B, HNF4A*),^13^ (2) within a commercially available MODY panel (*ABCC8, APPL1, BLK, EIF2AK3, FOXP3, GATA4, GATA6, GCK, GLIS3, HNF1A, HNF1B, HNF4A, IER3IP1, INS, KCNJ11, KLF11, MNX1, NEUROD1, NEUROG3, NKX2-2, PAX4, PDX1, PPARG, PTF1A, RFX6, SLC19A2, WFS1, ZFP57*),^19^ and (3) within a control panel comprising of genes from a commercially available hereditary cancer panel (**Supplemental Table 12**).

We assessed variant characteristics, including effect size and minor allele count, and generated density plots to assess the distribution of variant effect size stratified by pathogenicity classification category. We performed ANOVA testing and examined the means +/- the standard deviation to assess for differences in effect size between pathogenicity categories.We next assessed the estimated difference in individual endophenotype value for carriers of variants in each pathogenicity category compared to carriers of benign variants using a linear regression model. In the model, we regressed individual endophenotype values against carrier status of variants in each pathogenicity category, adjusting for age, sex, clinical covariates, and first 12 principal components of ancestry. Analyses were conducted for each of the gene panels of interest. We considered a two-sided P value of 0.05 significant. Analyses were performed using R version 3.6.^45^

### Sensitivity analyses in European ancestry cohorts

We performed sensitivity analyses in a UKBB European ancestry cohort for all analyses (LDL-C n=165,786, QTc n=28,249, HbA1c n=166,377). In single variant association testing, we used the first 4 principal components of ancestry; all other procedures remained the same.

### Replication analyses in FOURIER and TOPMed

Our LDL-C and HbA1c analyses was replicated in the Further Cardiovascular Outcomes Research With PCSK9 Inhibition in Subjects With Elevated Risk (FOURIER) trial cohort. We performed variant and sample QC as outlined above. The same clinical covariates were used; of note, all participants in FOURIER were taking statin medications. After sample and clinical trait exclusions as outlined above, 14,038 participants remained for our LDL-C replication analysis, and 12,798 participants remained for our HbA1c replication analysis in FOURIER.

Our QTc replication cohort included subjects from the National Heart Lung and Blood Institute’s (NHLBI) Trans-Omics for Precision Medicine (TOPMed) program^47^ with whole-genome sequencing (WGS) and ECG data. The present analysis includes nine studies including the Atherosclerosis Risk in Communities (ARIC) study, Genetics of Cardiometabolic Health in the Amish (Amish), Mount Sinai BioMe Biobank (BioMe), Cleveland Family Study (CFS), Cardiovascular Health Study (CHS), Framingham Heart Study (FHS), Jackson Heart Study (JHS), Multi-Ethnic Study of Atherosclerosis (MESA), and Women’s Health Initiative (WHI). Use of the TOPMed cohort for analysis was approved under paper proposal ID 8472. Genomic data included those available in Freeze 6. Details of comprising studies are provided in the supplement. Detailed WGS and variant calling protocols for the samples in TOPMed are provided on the TOPMed website.^15,47^ Briefly, 30X whole genome sequencing was performed, reads were aligned to human genome build GRCh38, joint genotype calling was performed, and initial sample QC was performed on all samples by the TOPMed Informatics Research Center. Clinical covariates including QT interval were defined using study-specific definitions. These measures were centrally collected and harmonized prior to our analysis.^48^ We performed variant and sample QC as outlined above. We subsetted the TOPMed WGS dataset to coding regions, defined as exons flanked by 5 base pairs, as previously described, for consistency with the UKBB exome cohort.^48^ After sample and clinical trait exclusions as outlined above, 26,976 participants remained for our replication analysis in TOPMed.

### Assessment of variant effect size as a discriminator for pathogenicity

We tested whether rare (MAF <1%) variant effect size discriminates pathogenic from non-pathogenic variants by fitting unadjusted logistic regression models in which we regressed the log-odds of a variant being pathogenic on the variant effect size. Only variants present in ClinVar were included in this analysis. Non-pathogenic variants included those adjudicated as likely benign and benign. In sensitivity analyses, we grouped pathogenic and likely pathogenic variants as one category and regressed on variant effect size. Analyses were conducted for each of the gene panels of interest in the primary and replication cohorts. We generated receiver operating characteristic (ROC) curves and calculated area under the curve (AUC) for each using the pROC package in R.^49^ We conducted a sensitivity analysis including only novel variants in the replication cohorts (FOURIER, TOPMed) not observed in the primary cohort (UKBB).

### Variant reclassification and nomination of potential pathogenic variants

Lastly, we used a high effect size threshold to provide evidence of pathogenicity for rare variants within genes of interest. We defined the “high effect size” threshold as effect size greater than 0.5 standard deviations (SD) of the trait distribution in the UKBB, the primary cohort. We applied this threshold to rare variants submitted to ClinVar with VUS and conflicting assertions and calculated the proportion of variants with evidence of pathogenicity using this method for each cohort. We also applied this threshold to rare variants not reported in ClinVar and calculated the proportion of variants with evidence of pathogenicity using this method for each cohort. Lastly, we tabulated the sensitivity and specificity of other effect size thresholds based on a range of SD cutoffs for each trait distribution.

## Data Availability

The data that support the findings of this study are available from the corresponding author upon reasonable request.

## CODE AVAILABILITY

Custom code or mathematical algorithms used to generate results reported in the manuscript can be obtained from contacting the corresponding author.

## ACKNOWLEDGMENTS

**UK Biobank**: Use of UK Biobank data was performed under application number 17488 and was approved by the local Massachusetts General Hospital institutional review board.

**Trans-Omics in Precision Medicine (TOPMed) program**: Molecular data for the Trans-Omics in Precision Medicine (TOPMed) program was supported by the National Heart, Lung and Blood Institute (NHLBI). Core support including read mapping and genotype calling, along with variant quality metrics and filtering were provided by the TOPMed Informatics Research Center (3R01HL-117626-02S1; contract HHSN268201800002I). Phenotype harmonization, data management, sample-identity QC, and general study coordination were provided by the TOPMed Data Coordinating Center (3R01HL-120393-02S1; contract HHSN268201800001I). We gratefully acknowledge the studies and participants who provided biological samples and data for TOPMed.

**Further Cardiovascular Outcomes Research With PCSK9 Inhibition in Subjects With Elevated Risk (FOURIER) clinical trial**: We wish to thank all participants on the FOURIER clinical trial. We would also like to acknowledge Giorgio Melloni and Nicholas Marston for facilitating our work with the FOURIER data. Clinical trial and exome sequencing was supported by Amgen.

