## Supplemental Material for "Endophenotype Effect Sizes Provide Evidence Supporting Variant Pathogenicity in Monogenic Disease Susceptibility Genes"

| **Items** | **Page** |
| --- | --- |
| Supplemental Methods | 3 |
| Supplemental Table 1. Baseline replication cohort characteristics by endophenotype | 4 |
| Supplemental Table 2. Baseline cohort characteristics by endophenotype in European-ancestry participants of the UK Biobank | 5 |
| Supplemental Table 3. Distribution and characteristics of variants in LDL-C associated genetic testing panels of interest | 6 |
| Supplemental Table 4. Distribution and characteristics of variants in QTc-associated genetic testing panels of interest | 7 |
| Supplemental Table 5. Distribution and characteristics of variants in MODY-associated genetic testing panels of interest | 8 |
| Supplemental Table 6. Distribution and characteristics of variants in LDL-C associated genetic testing panels of interest in FOURIER | 9 |
| Supplemental Table 7. Distribution and characteristics of variants in QTc-associated genetic testing panels of interest in TOPMed | 10 |
| Supplemental Table 8. Distribution and characteristics of variants in MODY-associated genetic testing panels of interest in FOURIER | 11 |
| Supplemental Table 9. Discriminative ability of various effect size thresholds | 12 |
| Supplemental Table 10. Characteristics of VUS and C variants with evidence of pathogenicity | See Excel |
| Supplemental Table 11. Characteristics of variants not reported in ClinVar with evidence of pathogenicity | See Excel |
| Supplemental Table 12. Genetic testing panels of interest | 13 |
| Supplemental Figure 1. CONSORT diagram of LDL-C endophenotype analysis | 14 |
| Supplemental Figure 2. CONSORT diagram of QTc endophenotype analysis | 15 |
| Supplemental Figure 3. CONSORT diagram of MODY endophenotype analysis | 16 |
| Supplemental Figure 4. Association between variant effect size and pathogenicity for three endophenotypes and a control panel | 17 |
| Supplemental Figure 5. Association between variant effect size and pathogenicity for three endophenotypes in a European-ancestry cohort | 18 |
| Supplemental Figure 6. Association between variant effect size and pathogenicity for three endophenotypes for variants with MAC ≤ 5 | 19 |
| Supplemental Figure 7. Association between variant effect size and pathogenicity for three endophenotypes in replication cohorts | 20 |
| Supplemental Figure 8. Discrimination of variant pathogenicity by effect size with replication variants independent of the UK Biobank | 21 |
| Supplemental Acknowledgements | 22-25 |
| Supplemental References | 26 |

**Supplemental Methods**

*Estimation of genetic ancestry for TOPMed*

To estimate genetic ancestry within the TOPMed cohort, we selected common (MAF ≥ 5%), high confidence (missingness rate <1%) variants in the 1000G dataset that were also present in TOPMed. We pruned these variants within five ethnic groups in the 1000G dataset (European, East Asian, South Asian, Admixed American, and African American) using PLINK (“--indep-pairwise 50 5 0.2).^1^ We used ADMIXTURE to learn the genetic structure of the five reference ethnic groups, and then projected the TOPMed participants on the reference groups.^2^ We classified TOPMed individuals into one of the five ethnic groups when the probability of being a member of that group was > 80%. Individuals with <80% probability of being a member of all five ethnic groups were classified as “Undetermined”.

*Clinical covariates and exclusion criteria in the UKBB*

Baseline history of myocardial infarction was defined centrally using an algorithmically defined outcome, which takes self-reports, hospital admissions and death records into account.^3^ Baseline history of heart failure was defined using: self-reported non-cancer illness codes 1076, 1079 and 1588; ICD10 codes I11.0, I13.0, I.13.2, I25.5, I42.0, I42.1, I42.2, I42.5, I42.8, I42.9, I50, I50.0, I50.1 and I50.9 among main diagnoses, secondary diagnoses, primary causes of death and secondary causes of death; and ICD9 codes 4254, 4280, 4281 and 4289 among main and secondary diagnoses. Presence of cardiac pacemaker was defined using: self-reported presence of pacemaker during interview; self-reported operation codes 1096, 1548 and 1549; ICD10 code Z45.0 among main and secondary diagnoses; operative procedure codes K60, K60.1, K60.2, K60,3, K60.4, K60.5, K60.6, K60.7, K60.8, K60.9, K61, K61.1, K61.2, K61.3, K61.4, K61.5, K61.6, K61.7, K61.8 and K61.9 among main and secondary OPCS. Wolff-Parkinson-White syndrome was defined using: self-reported non-cancer illness code 1484; ICD10 code I45.6 among main diagnoses, secondary diagnoses, primary causes of death and secondary causes of death; ICD9 code 4267 among primary and secondary diagnoses; operative procedure codes K52.4 and K57.4 among main and secondary OPCS. Additionally, for the LDL analysis, HDL was measured by the same Beckman Coulter AU5800 device at the time of LDL measurement. Statin usage was ascertained via self-reported data at initial assessment.

### **Supplemental Table 1.** Baseline replication cohort characteristics by endophenotype

| **Characteristic** | **LDL-C (mg/dL)** | **QTc (ms)** | **HbA1c (%)** |
| --- | --- | --- | --- |
| Cohort name | FOURIER | TOPMed | FOURIER |
| Participants with a measurable endophenotype, n | 14,038 | 26,976 | 12,798 |
| Median endophenotype value (Q1-Q3) | 92.0 (80.0-109.0) | 421.0 (409.0-437.0) | 5.8 (5.5-6.3) |
| Male, n (%) | 10,720 (76.4) | 9,332 (34.6) | 9,731 (76.0) |
| European ancestry, n (%) | 14,023 (100.0%) | 16,074 (59.5) | 12,783 (99.8) |
| Mean age, years (SD) | 62.8 (8.8) | 59.8 (12.5) | 62.9 (8.8) |
| Myocardial infarction, n (%) | 11,470 (81.7) | 2,361 (8.8) | - |
| Statin usage, n (%) | 14,039 (100.0) | - | - |
| Mean high-density lipoprotein, mg/dL (SD) | 46.7 (12.7) | - | - |
| Heart failure, n (%) | - | 1,788 (6.6) | - |
| Beta blocker usage, n (%) | - | 3,415 (12.6) | - |
| Calcium channel blocker usage, n (%) | - | 3,043 (11.3) | - |
| Type 2 diabetes medication usage, n (%) | - | - | 3,265 (25.6) |
| Mean corpuscular volume, femtoliters (SD) | - | - | 92.8 (4.8) |
| NB: only select relevant characteristics for the given monogenic disease of interest are displayed | | | |

**Supplemental Table 2**. Baseline cohort characteristics by endophenotype in European-ancestry participants of the UK Biobank

| **Characteristic** | **LDL-C (mg/dL)** | **QTc (ms)** | **HbA1c (%)** |
| --- | --- | --- | --- |
| Participants with a measurable endophenotype, n | 165,786 | 28,249 | 166,337 |
| Median endophenotype value (Q1-Q3) | 136.6 (114.5-159.6) | 411.3 (396.4-426.4) | 5.4 (5.1-5.6) |
| Male, n (%) | 74,749 (45.1) | 14,138 (50.0) | 74,984 (45.1) |
| Mean age, years (SD) | 57.3 (8.0) | 53.0 (5.6) | 57.3 (8.0) |
| Myocardial infarction, n (%) | 2,668 (1.6) | 422 (1.5) | - |
| Statin usage, n (%) | 27,796 (16.8) | - | - |
| Mean high-density lipoprotein, mg/dL (SD) | 56.3 (14.2) | - | - |
| Heart failure, n (%) | - | 66 (0.2) | - |
| Beta blocker usage, n (%) | - | 1,490 (5.3) | - |
| Calcium channel blocker usage, n (%) | - | 1,968 (7.0) | - |
| Type 2 diabetes medication usage, n (%) | - | - | 5,700 (3.4) |
| Mean corpuscular volume, femtoliters (SD) | - | - | 91.4 (4.2) |
| NB: only select relevant characteristics for the given monogenic disease of interest are displayed | | | |

**Supplemental Table 3**. Distribution and characteristics of variants in LDL-associated genetic testing panels of interest

|  | **Definitive FH genes** | **Commercially available**  **FH panel** | **Commercially available hereditary cancer panel** |
| --- | --- | --- | --- |
| Total variants in SVA | 3,505 | 3,676 | 25,078 |
| Variants of MAF < 1% | 3,487 | 3,655 | 24,953 |
| Variants of MAF < 1% not in ClinVar | 2,030 | 2,159 | 7,656 |
| Variants of MAF < 1% in ClinVar | 1,457 | 1,496 | 17,297 |
| Pathogenic (P) variants | 44 | 46 | 755 |
| Mean effect size, mg/dL(SD) | 47.02 (53.10) | 44.91 (52.86) | -0.54 (24.60) |
| Median MAC (Q1-Q3) | 2 (1-5) | 2 (1-4.75) | 2 (1-4) |
| Likely pathogenic (LP) variants | 18 | 18 | 163 |
| Mean effect size, mg/dL(SD) | 44.50 (58.43) | 44.50 (58.43) | -2.52 (24.14) |
| Median MAC (Q1-Q3) | 2 (1-2) | 2 (1-2) | 2 (1-4) |
| Likely benign (LB) variants | 410 | 422 | 4,350 |
| Mean effect size, mg/dL(SD) | -2.15 (21.52) | -2.04 (21.36) | 0.25 (21.86) |
| Median MAC (Q1-Q3) | 4 (2-10) | 4 (2-10.75) | 2 (1-6) |
| Benign (B) variants | 45 | 45 | 1,014 |
| Mean effect size, mg/dL(SD) | -0.95 (12.69) | -0.95 (12.69) | 0.05 (11.59) |
| Median MAC (Q1-Q3) | 22 (7-55) | 22 (7-55) | 28 (8-101) |
| Variants of uncertain significance (VUS) | 620 | 638 | 8,693 |
| Mean effect size, mg/dL(SD) | -2.06 (22.01) | -1.98 (22.00) | 0.56 (23.30) |
| Median MAC (Q1-Q3) | 4 (2-10) | 4 (2-10) | 2 (1-4) |
| Variants of conflicting interpretations (C) | 320 | 327 | 2,322 |
| Mean effect size, mg/dL(SD) | 4.45 (19.80) | 4.36 (19.60) | -0.05 (16.22) |
| Median MAC (Q1-Q3) | 14 (4-67.5) | 14 (4-71) | 7 (2-22) |

### FH: Familial Hypercholesterolemia, SVA: single variant association, MAF: minor allele frequency, B: benign, LB: likely benign, LP: likely pathogenic, P: pathogenic, VUS: variant of uncertain significance, C: conflicting

**Supplemental Table 4**. Distribution and characteristics of variants in QTc-associated genetic testing panels of interest

|  | **Definitive LQTS genes** | **Commercially available**  **LQTS panel** | **Commercially available hereditary cancer panel** |
| --- | --- | --- | --- |
| Total variants in SVA | 1,091 | 2,217 | 11,450 |
| Variants of MAF < 1% | 1,083 | 2,201 | 11,316 |
| Variants of MAF < 1% not in ClinVar | 319 | 673 | 2,782 |
| Variants of MAF < 1% in ClinVar | 764 | 1,528 | 8,534 |
| Pathogenic (P) variants | 19 | 23 | 287 |
| Mean effect size, ms (SD) | 30.10 (29.90) | 26.79 (28.10) | 0.89 (19.57) |
| Median MAC (Q1-Q3) | 1 (1-2) | 2 (1-2) | 1 (1-2) |
| Likely pathogenic (LP) variants | 13 | 16 | 64 |
| Mean effect size, ms (SD) | 11.54 (36.98) | 11.01 (33.55) | -1.64 (26.68) |
| Median MAC (Q1-Q3) | 2 (1-2) | 1.5 (1-2) | 1 (1-2) |
| Likely benign (LB) variants | 230 | 470 | 2,126 |
| Mean effect size, ms (SD) | 1.80 (17.00) | 1.84 (17.31) | 0.60 (19.23) |
| Median MAC (Q1-Q3) | 2 (1-4) | 2 (1-4) | 1 (1-3) |
| Benign (B) variants | 47 | 146 | 869 |
| Mean effect size, ms (SD) | -0.58 (11.10) | 0.51 (8.85) | -0.04 (12.02) |
| Median MAC (Q1-Q3) | 8 (4-26) | 13 (5-47.75) | 9 (3-31) |
| Variants of uncertain significance (VUS) | 276 | 574 | 3,644 |
| Mean effect size, ms (SD) | 0.08 (18.46) | 0.55 (18.00) | -0.47 (19.29) |
| Median MAC (Q1-Q3) | 1 (1-3) | 2 (1-3) | 1 (1-2) |
| Variants of conflicting interpretations (C) | 179 | 299 | 1,544 |
| Mean effect size, ms (SD) | 0.89 (15.67) | 0.67 (13.51) | -0.08 (15.69) |
| Median MAC (Q1-Q3) | 3 (1.5-11) | 4 (2-15) | 3 (1-8) |

### LQTS: Long-QT syndrome, SVA: single variant association, MAF: minor allele frequency, B: benign, LB: likely benign, LP: likely pathogenic, P: pathogenic, VUS: variant of uncertain significance, C: conflicting

**Supplemental Table 5.** Distribution and characteristics of variants in MODY-associated genetic testing panels of interest

|  | **Common MODY genes** | **Commercially available**  **MODY panel** | **Commercially available hereditary cancer panel** |
| --- | --- | --- | --- |
| Total variants in SVA | 1,052 | 6,600 | 24,962 |
| Variants of MAF < 1% | 1,046 | 6,565 | 24,836 |
| Variants of MAF < 1% not in ClinVar | 859 | 5,723 | 7,591 |
| Variants of MAF < 1% in ClinVar | 187 | 842 | 17,245 |
| Pathogenic (P) variants | 20 | 52 | 754 |
| Mean effect size, % (SD) | 0.70 (0.70) | 0.31 (0.56) | 0.01 (0.38) |
| Median MAC (Q1-Q3) | 1.5 (1-2) | 2 (1-3) | 2 (1-4) |
| Likely pathogenic (LP) variants | 6 | 24 | 161 |
| Mean effect size, % (SD) | 0.45 (0.37) | 0.25 (0.50) | -0.03 (0.28) |
| Median MAC (Q1-Q3) | 1 (1-1.75) | 1 (1-2) | 2 (1-4) |
| Likely benign (LB) variants | 29 | 188 | 4,311 |
| Mean effect size, % (SD) | -0.05 (0.19) | 0.00 (0.35) | -0.01 (0.39) |
| Median MAC (Q1-Q3) | 11 (4-33) | 8.5 (3-24) | 2 (1-7) |
| Benign (B) variants | 30 | 132 | 1,019 |
| Mean effect size, % (SD) | 0.05 (0.20) | 0.00 (0.14) | 0.00 (0.20) |
| Median MAC (Q1-Q3) | 39 (16-76) | 71 (24.5-222.5) | 27 (7-95) |
| Variants of uncertain significance (VUS) | 67 | 301 | 8,678 |
| Mean effect size, % (SD) | 0.01 (0.27) | 0.02 (0.28) | -0.00 (0.43) |
| Median MAC (Q1-Q3) | 5 (2-17.5) | 6 (2-22) | 2 (1-4) |
| Variants of conflicting interpretations (C) | 35 | 145 | 2,322 |
| Mean effect size, % (SD) | 0.04 (0.23) | 0.01 (0.17) | 0.00 (0.33) |
| Median MAC (Q1-Q3) | 11 (5.5-39.5) | 21 (8-83) | 7 (2-21) |

### MODY: maturity-onset diabetes of the young, SVA: single variant association, MAF: minor allele frequency, B: benign, LB: likely benign, LP: likely pathogenic, P: pathogenic, VUS: variant of uncertain significance, C: conflicting

**Supplemental Table 6**. Distribution and characteristics of variants in LDL-associated genetic testing panels of interest in FOURIER

|  | **Definitive FH genes** | **Commercially available**  **FH panel** | **Commercially available hereditary cancer panel** |
| --- | --- | --- | --- |
| Total variants in SVA | 898 | 938 | 5,402 |
| Variants of MAF < 1% | 883 | 921 | 5,296 |
| Variants of MAF < 1% not in ClinVar | 324 | 346 | 892 |
| Variants of MAF < 1% in ClinVar | 559 | 575 | 4,404 |
| Pathogenic (P) variants | 58 | 59 | 145 |
| Mean effect size, mg/dL(SD) | 61.19 (58.15) | 61.00 (57.66) | 4.09 (30.16) |
| Median MAC (Q1-Q3) | 1 (1-2) | 1 (1-1.5) | 1 (1-2) |
| Likely pathogenic (LP) variants | 19 | 19 | 33 |
| Mean effect size, mg/dL(SD) | 36.70 (57.75) | 36.7 (57.7) | -0.31 (19.45) |
| Median MAC (Q1-Q3) | 2 (1-2) | 1 (1-1.5) | 1 (1-1) |
| Likely benign (LB) variants | 102 | 104 | 988 |
| Mean effect size, mg/dL(SD) | 2.69 (21.87) | 2.67 (21.90) | -0.40 (23.35) |
| Median MAC (Q1-Q3) | 2 (1-5) | 1 (1-2) | 1 (1-2) |
| Benign (B) variants | 19 | 19 | 524 |
| Mean effect size, mg/dL(SD) | -4.95 (12.89) | -4.95 (12.89) | 1.21 (18.86) |
| Median MAC (Q1-Q3) | 2 (1-5) | 1 (1-2) | 3 (1-10) |
| Variants of uncertain significance (VUS) | 192 | 199 | 1827 |
| Mean effect size, mg/dL(SD) | -0.25 (22.98) | -0.52 (22.72) | 0.90 (30.69) |
| Median MAC (Q1-Q3) | 1 (1-2) | 1 (1-2) | 1 (1-1) |
| Variants of conflicting interpretations (C) | 169 | 175 | 887 |
| Mean effect size, mg/dL(SD) | 8.51 (44.94) | 8.07 (44.25) | -0.11 (18.83) |
| Median MAC (Q1-Q3) | 2 (1-8) | 3 (1-9) | 2 (1-5) |

### FH: Familial Hypercholesterolemia, SVA: single variant association, MAF: minor allele frequency, B: benign, LB: likely benign, LP: likely pathogenic, P: pathogenic, VUS: variant of uncertain significance, C: conflicting

**Supplemental Table 7.** Distribution and characteristics of variants in QTc-associated genetic testing panels of interest in TOPMed

|  | **Definitive LQTS genes** | **Commercially available**  **LQTS panel** | **Commercially available hereditary cancer panel** |
| --- | --- | --- | --- |
| Total variants in SVA | 1,190 | 2,471 | 11,915 |
| Variants of MAF < 1% | 1,171 | 2,427 | 11,697 |
| Variants of MAF < 1% not in ClinVar | 327 | 769 | 2,144 |
| Variants of MAF < 1% in ClinVar | 844 | 1,658 | 9,553 |
| Pathogenic (P) variants | 23 | 30 | 282 |
| Mean effect size, ms (SD) | 37.20 (35.94) | 29.54 (35.14) | 2.67 (20.93 |
| Median MAC (Q1-Q3) | 1 (1-1.5) | 1 (1-2) | 1 (1-2) |
| Likely pathogenic (LP) variants | 12 | 15 | 65 |
| Mean effect size, ms (SD) | 11.61 (34.24) | 11.53 (30.54) | 0.75 (20.32) |
| Median MAC (Q1-Q3) | 1 (1-2.25) | 1 (1-2) | 1 (1-2) |
| Likely benign (LB) variants | 243 | 499 | 2,398 |
| Mean effect size, ms (SD) | 1.48 (25.19) | 1.19 (20.87) | 0.38 (18.52) |
| Median MAC (Q1-Q3) | 2 (1-3) | 2 (2-5) | 1 (1-3) |
| Benign (B) variants | 49 | 146 | 917 |
| Mean effect size, ms (SD) | 1.89 (11.67) | 0.86 (8.65) | 0.10 (10.80) |
| Median MAC (Q1-Q3) | 24 (3-75) | 42 (6.25-148.25) | 13 (1-89) |
| Variants of uncertain significance (VUS) | 303 | 636 | 4,145 |
| Mean effect size, ms (SD) | -0.86 (21.29) | -0.86 (18.95) | -0.33 (19.70) |
| Median MAC (Q1-Q3) | 1 (1-3) | 2 (1-3) | 1 (1-2) |
| Variants of conflicting interpretations (C) | 214 | 332 | 1,746 |
| Mean effect size, ms (SD) | 0.81 (14.83) | -0.27 (13.42) | 0.51 (15.37) |
| Median MAC (Q1-Q3) | 3 (2-11.75) | 4 (2-17) | 3 (1-8) |

### LQTS: Long-QT syndrome, SVA: single variant association, MAF: minor allele frequency, B: benign, LB: likely benign, LP: likely pathogenic, P: pathogenic, VUS: variant of uncertain significance, C: conflicting

**Supplemental Table 8**. Distribution and characteristics of variants in MODY-associated genetic testing panels of interest in FOURIER

|  | **Common MODY genes** | **Commercially available**  **MODY panel** | **Commercially available hereditary cancer panel** |
| --- | --- | --- | --- |
| Total variants in SVA | 387 | 2,370 | 9,993 |
| Variants of MAF < 1% | 375 | 2,326 | 9,781 |
| Variants of MAF < 1% not in ClinVar | 320 | 2,017 | 5,640 |
| Variants of MAF < 1% in ClinVar | 55 | 309 | 4,141 |
| Pathogenic (P) variants | 2 | 10 | 134 |
| Mean effect size, ms (SD) | 1.68 (0.55) | 0.41 (1.01) | 0.08 (0.88) |
| Median MAC (Q1-Q3) | 1 (1-1) | 1 (1-1) | 1 (1-1) |
| Likely pathogenic (LP) variants | 4 | 5 | 30 |
| Mean effect size, ms (SD) | -0.06 (1.04) | 0.05 (0.94) | 0.18 (0.79) |
| Median MAC (Q1-Q3) | 1 (1-1) | 1 (1-1) | 1 (1-1) |
| Likely benign (LB) variants | 11 | 65 | 927 |
| Mean effect size, ms (SD) | 0.32 (0.98) | 0.04 (0.78) | 0.02 (0.86) |
| Median MAC (Q1-Q3) | 3 (1-5) | 2 (1-7) | 1 (1-2) |
| Benign (B) variants | 7 | 61 | 508 |
| Mean effect size, ms (SD) | -0.14 (0.31) | 0.04 (0.53) | 0.01 (0.63) |
| Median MAC (Q1-Q3) | 2 (1-3) | 5 (2-18) | 3 (1-10) |
| Variants of uncertain significance (VUS) | 17 | 97 | 1,693 |
| Mean effect size, ms (SD) | 0.09 (0.94) | -0.05 (0.71) | 0.02 (0.86) |
| Median MAC (Q1-Q3) | 2 (1-4) | 1 (1-3) | 1 (1-1) |
| Variants of conflicting interpretations (C) | 14 | 71 | 849 |
| Mean effect size, ms (SD) | -0.17 (0.69) | -0.06 (0.46) | 0.01 (0.68) |
| Median MAC (Q1-Q3) | 1.5 (1-8.75) | 5 (1-15) | 2 (1-5) |

### MODY: maturity-onset diabetes of the young, SVA: single variant association, MAF: minor allele frequency, B: benign, LB: likely benign, LP: likely pathogenic, P: pathogenic, VUS: variant of uncertain significance, C: conflicting

**Supplemental Table 9.** Discriminative ability of various effect size thresholds

| Endophenotype | **LDL-C** | | | **QTc** | | | **HbA1c** | | |
| --- | --- | --- | --- | --- | --- | --- | --- | --- | --- |
| Standard deviation of endophenotype distribution | Effect size (mg/dL) | Sensitivity^#^ (%) | Specificity^#^ (%) | Effect size (ms) | Sensitivity (%) | Specificity (%) | Effect size (%) | Sensitivity (%) | Specificity (%) |
| 0.125 | 4.18 | 82 | 67 | 2.97 | 84 | 61 | 0.08 | 80 | 80 |
| 0.25 | 8.36 | 80 | 75 | 5.93 | 84 | 67 | 0.15 | 80 | 88 |
| 0.50* | 16.72 | 75 | 87 | 11.86 | 79 | 75 | 0.31 | 70 | 95 |
| 0.75 | 25.09 | 68 | 95 | 17.80 | 74 | 85 | 0.46 | 65 | 98 |
| 1.00 | 33.45 | 66 | 97 | 23.73 | 58 | 92 | 0.61 | 65 | 98 |

**A priori* determined threshold

^#^Sensitivity and specificity are properties of the effect size threshold’s ability to discriminate ClinVar designated pathogenic variants from non-pathogenic variants (likely benign and benign).

### **Supplemental Table 12**. Genetic testing panels of interest

| **Gene panel name** | **Number of genes** | **Genes included** |
| --- | --- | --- |
| Definitive FH genes | 3 | *LDLR, APOB, PCSK9* |
| Commercially available FH panel | 4 | *LDLR, APOB, PCSK9,* *LDLRAP* |
| Definitive LQTS genes | 3 | *KCNQ1, KCNH2, SCN5A* |
| Commercially available LQTS panel | 13 | *ANK2, CACNA1C, CALM1, CALM2, CALM3, CAV3*^#^, KCNE1*^#^, KCNE2*^#^, KCNH2, KCNJ2*^#^, KCNQ1, SCN5A, TRDN* |
| Common MODY genes | 4 | *GCK, HNF1A, HNF1B, HNF4A* |
| Commercially available MODY panel | 28 | *ABCC8, APPL1, BLK, EIF2AK3, FOXP3^#^, GATA4, GATA6, GCK, GLIS3, HNF1A, HNF1B, HNF4A, IER3IP1, INS, KCNJ11, KLF11, MNX1^#^, NEUROD1^#^, NEUROG3^#^, NKX2-2^#^, PAX4, PDX1^#^, PPARG, PTF1A, RFX6, SLC19A2, WFS1, ZFP57* |
| Commercially available hereditary cancer panel (control) | 47 | *APC, ATM, AXIN2, BARD1, BMPR1A, BRCA1, BRCA2, BRIP1, CDH1, CDK4, CDKN2A, CHEK2, CTNNA1, DICER1, EPCAM, GREM1*^#^, HOXB13*^#^, KIT, MEN1, MLH1, MSH2, MSH3, MSH6, MUTYH, NBN, NF1, NTHL1, PALB2, PDGFRA, PMS2, POLD1, POLE, PTEN, RAD50, RAD51C, RAD51D, SDHA, SDHB, SDHC, SDHD, SMAD4, SMARCA4, STK11, TP53, TSC1, TSC2, VHL* |
| ^#^No variants from ClinVar were recovered from this gene in UK Biobank analyses  *No variants from ClinVar were recovered from this gene in replication analyses | | |

FH: Familial hypercholesterolemia

LQTS: Long-QT syndrome

MODY: Maturity-onset diabetes of the young

**Supplemental Figure 1.** CONSORT diagram of LDL-C endophenotype analysis

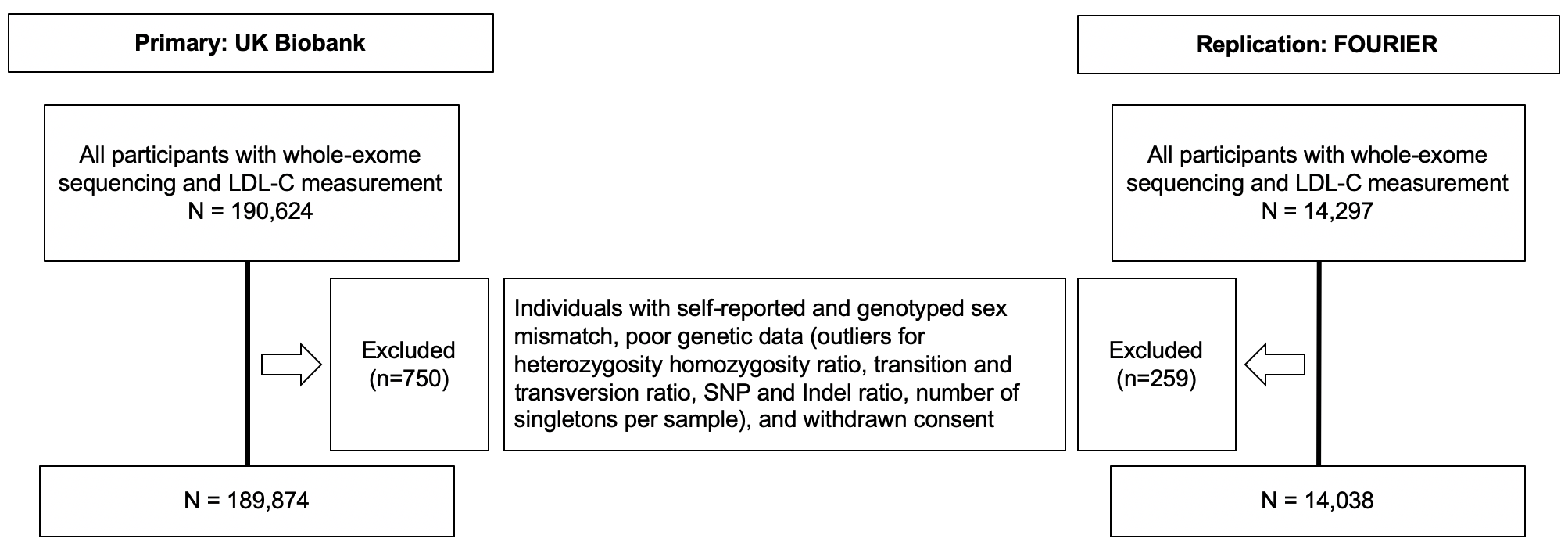

Each row from top to bottom shows the numbers of individuals excluded at each step. The individuals excluded from the UKBB cohort are on the left, and from the FOURIER cohort are on the right.

**Supplemental Figure 2**. CONSORT diagram of QTc endophenotype analysis

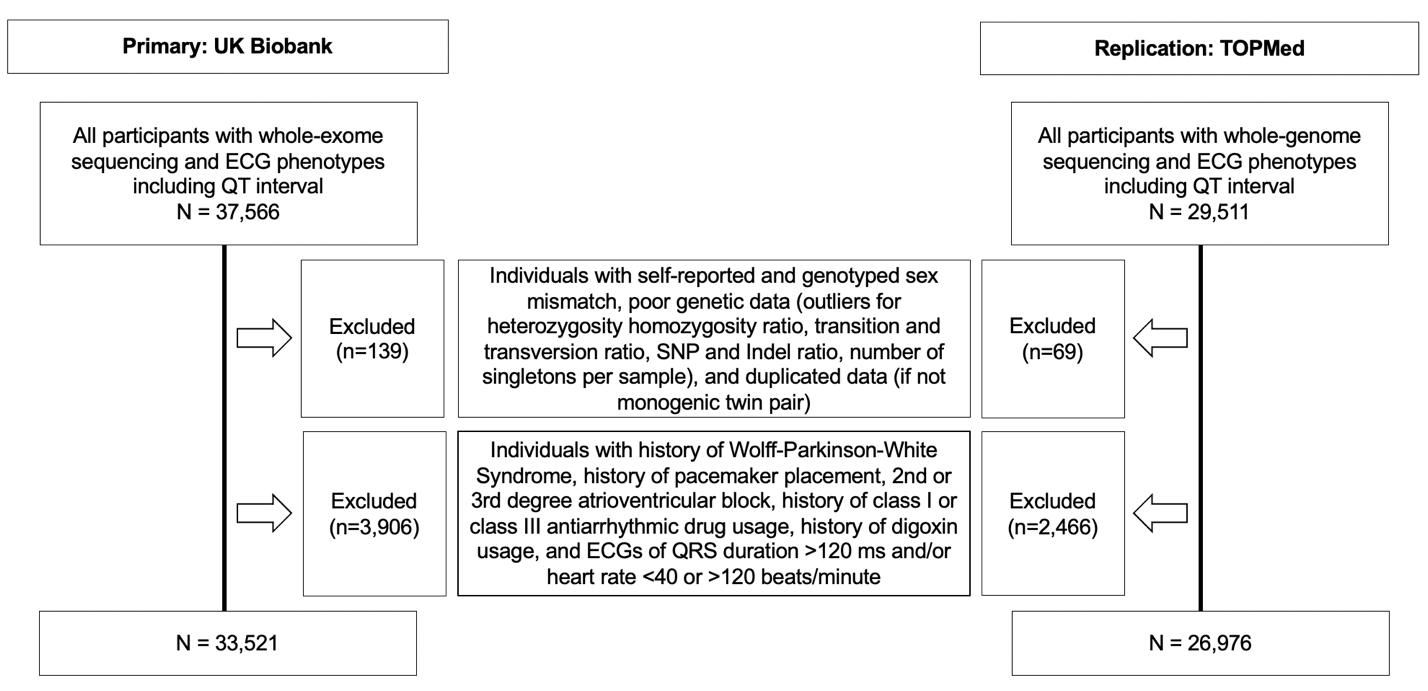

Each row from top to bottom shows the numbers of individuals excluded at each step. The individuals excluded from the UKBB cohort are on the left, and from the TOPMed cohort are on the right.

**Supplemental Figure 3.** CONSORT diagram of MODY endophenotype analysis

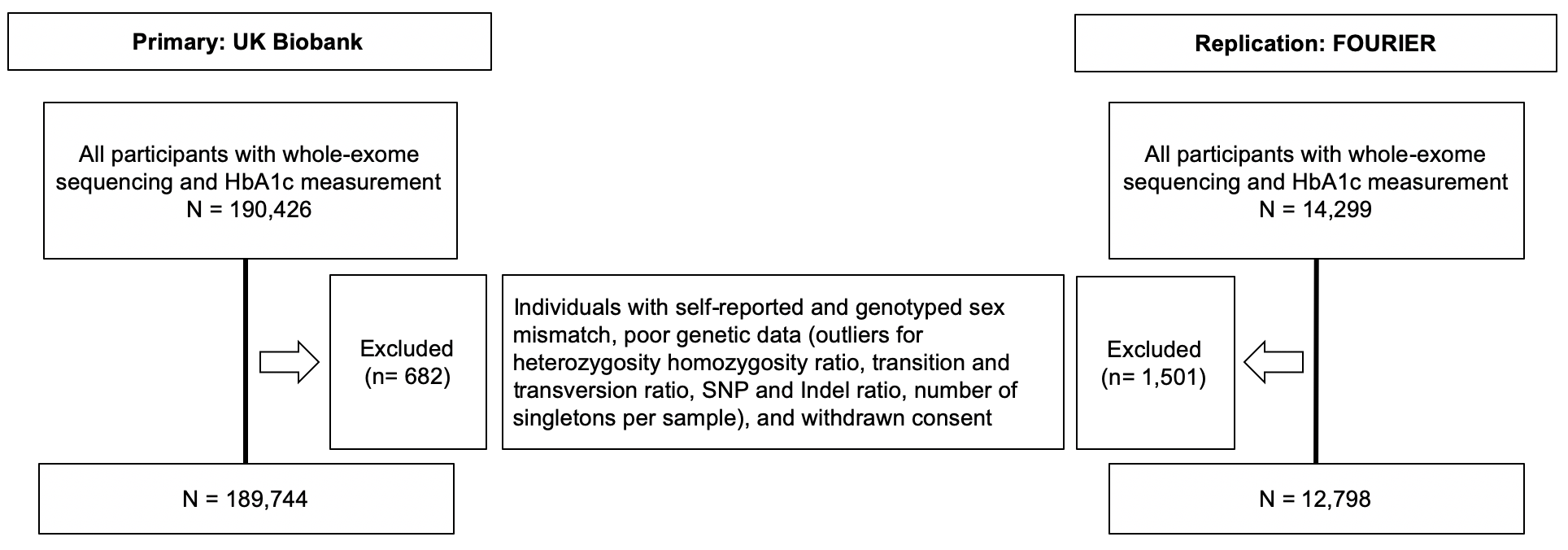

Each row from top to bottom shows the numbers of individuals excluded at each step. The individuals excluded from the UKBB cohort are on the left, and from the FOURIER cohort are on the right.

**Supplemental Figure 4.** Association between variant effect size and pathogenicity for three endophenotypes and a control panel

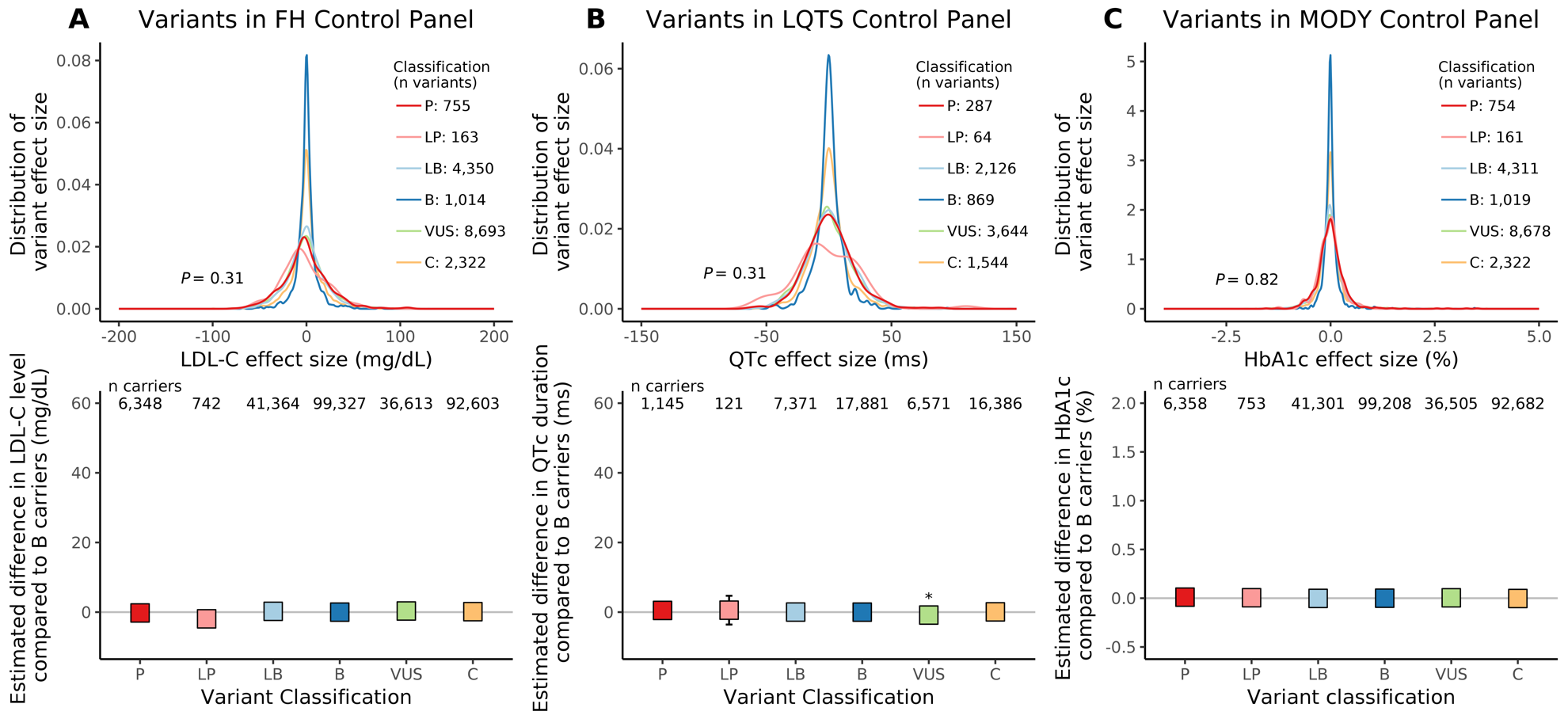
Association between effect size and variant pathogenicity for three monogenic disease endophenotypes in the control panel of hereditary cancer genes. Panels A, B, and C display data for the LDL-C, QTc, and HbA1c endophenotypes, respectively, for rare variants found in the UK Biobank. Row 1 in each panel displays the variant effect size distribution by ClinVar pathogenicity category (colored). Row 2 in each panel displays the estimated difference in endophenotype value comparing carriers of a variant in each ClinVar pathogenicity category to carriers of benign variants (boxes) and 95% confidence intervals (whiskers); * indicates *P* < 0.05. Variant classification includes B: benign, LB: likely benign, LP: likely pathogenic, P: pathogenic, VUS: variant of uncertain significance, C: conflicting.

**Supplemental Figure 5**. Association between variant effect size and pathogenicity for three endophenotypes in a European-ancestry cohort

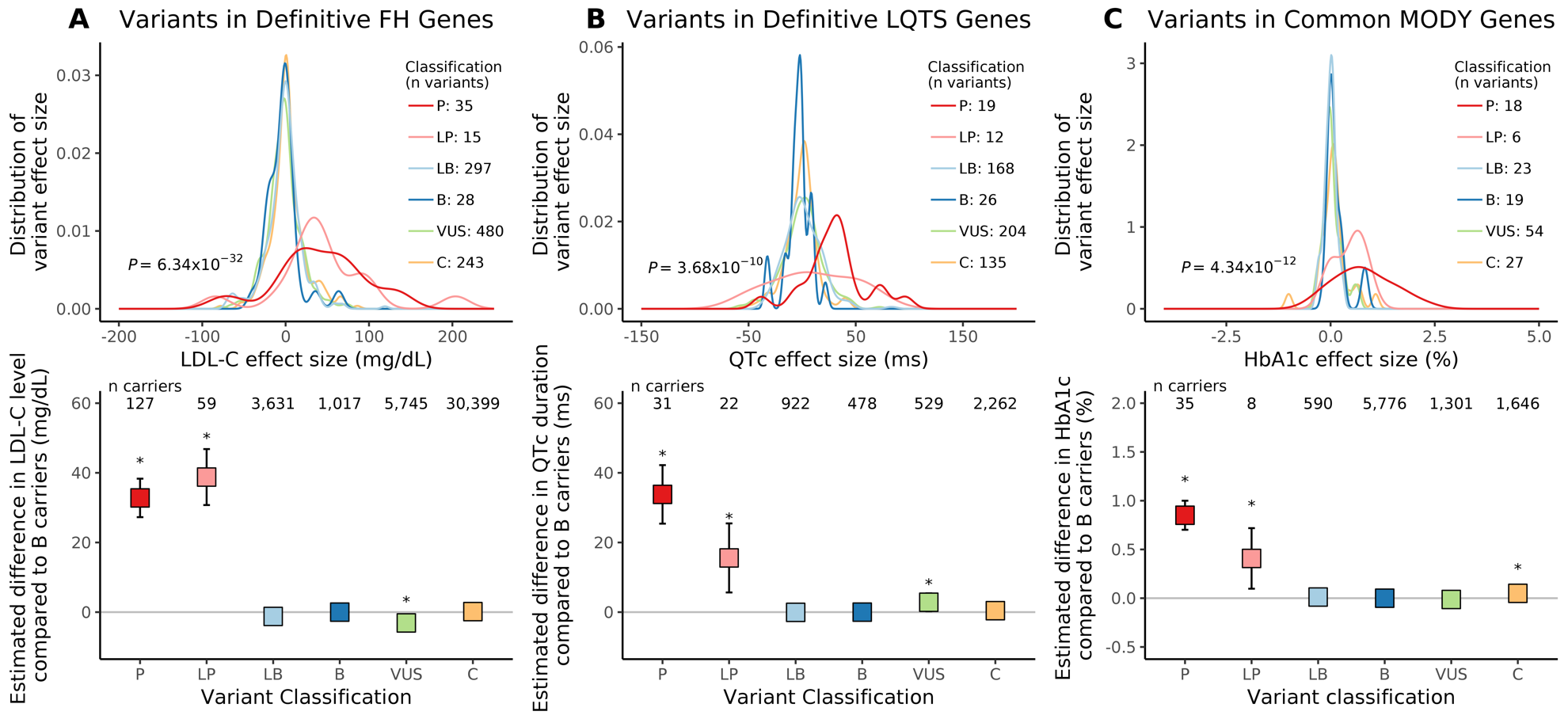
Association between effect size and variant pathogenicity for three monogenic disease endophenotypes for rare variants found in participants of white European ancestry in the UK Biobank. Panels A, B, and C display data for the LDL-C, QTc, and HbA1c endophenotypes, respectively, for rare variants found in the UK Biobank. Definitive familial hypercholesterolemia (FH) genes include *LDLR, APOB, PCSK9*; definitive long-QT syndrome (LQTS) genes include *KCNQ1, KCNH2, SCN5A*; common maturity-onset diabetes of the young (MODY) genes include *HNF1A, HNF1B, HNF4A, GCK*. Row 1 in each panel displays the variant effect size distribution by ClinVar pathogenicity category (colored). Row 2 in each panel displays the estimated difference in endophenotype value comparing carriers of a variant in each ClinVar pathogenicity category to carriers of benign variants (boxes) and 95% confidence intervals (whiskers); * indicates *P* < 0.05. Variant classification includes B: benign, LB: likely benign, LP: likely pathogenic, P: pathogenic, VUS: variant of uncertain significance, C: conflicting.

### **Supplemental Figure 6.** Association between variant effect size and pathogenicity for three endophenotypes for variants with MAC ≤ 5

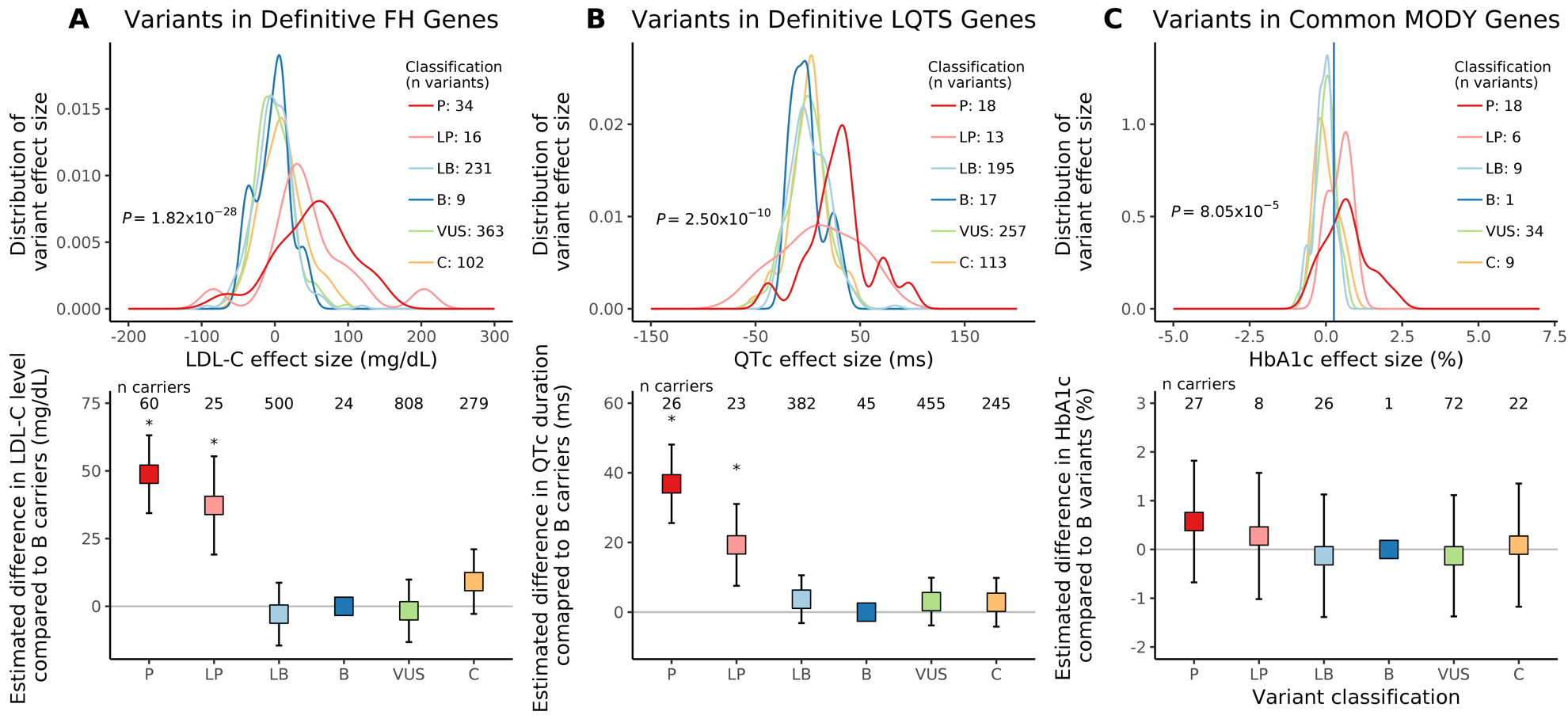
Association between effect size and variant pathogenicity for three monogenic disease endophenotypes, for variants with minor allele count ≤ 5 in the UK Biobank. Panels A, B, and C display data for the LDL-C, QTc, and HbA1c endophenotypes, respectively, for rare variants found in the UK Biobank. Definitive familial hypercholesterolemia (FH) genes include *LDLR, APOB, PCSK9*; definitive long-QT syndrome (LQTS) genes include *KCNQ1, KCNH2, SCN5A*; common maturity-onset diabetes of the young (MODY) genes include *HNF1A, HNF1B, HNF4A, GCK*. Row 1 in each panel displays the variant effect size distribution by ClinVar pathogenicity category (colored). Row 2 in each panel displays the estimated difference in endophenotype value comparing carriers of a variant in each ClinVar pathogenicity category to carriers of benign variants (boxes) and 95% confidence intervals (whiskers); * indicates *P* < 0.05. Variant classification includes B: benign, LB: likely benign, LP: likely pathogenic, P: pathogenic, VUS: variant of uncertain significance, C: conflicting.

**Supplemental Figure 7**. Association between variant effect size and pathogenicity for three endophenotypes in replication cohorts

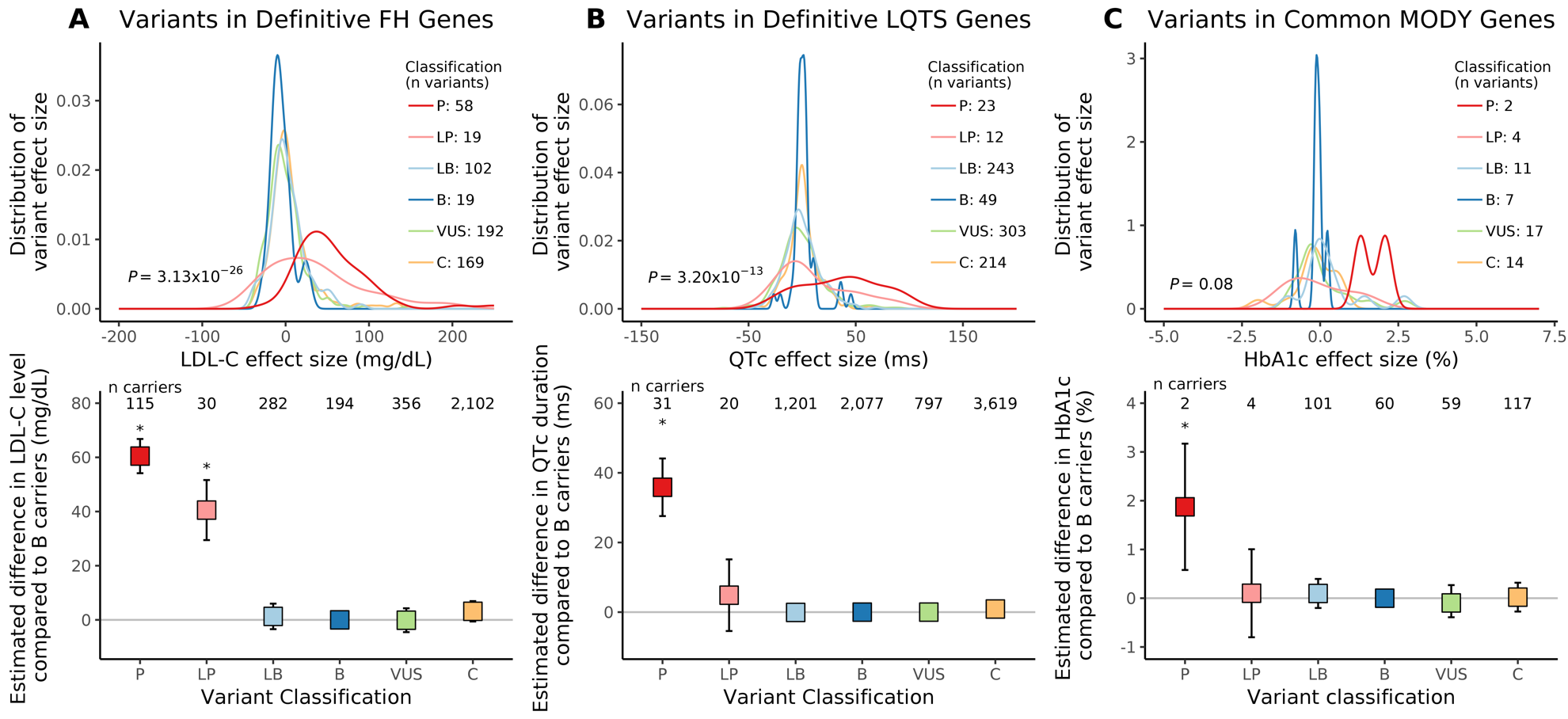
Association between effect size and variant pathogenicity for three monogenic disease endophenotypes, for rare variants found in replication cohorts. Panels A, B, and C display data for the LDL-C, QTc, and HbA1c endophenotypes, respectively, for rare variants found in the FOURIER, TOPMed, and FOURIER, respectively. Definitive familial hypercholesterolemia (FH) genes include *LDLR, APOB, PCSK9*; definitive long-QT syndrome (LQTS) genes include *KCNQ1, KCNH2, SCN5A*; common maturity-onset diabetes of the young (MODY) genes include *HNF1A, HNF1B, HNF4A, GCK*. Row 1 in each panel displays the variant effect size distribution by ClinVar pathogenicity category (colored). Row 2 in each panel displays the estimated difference in endophenotype value comparing carriers of a variant in each ClinVar pathogenicity category to carriers of benign variants (boxes) and 95% confidence intervals (whiskers); * indicates *P* < 0.05. Variant classification includes B: benign, LB: likely benign, LP: likely pathogenic, P: pathogenic, VUS: variant of uncertain significance, C: conflicting.

### **Supplemental Figure 8.** Discrimination of variant pathogenicity by effect size with novel variants in replication cohorts not observed in the UK Biobank

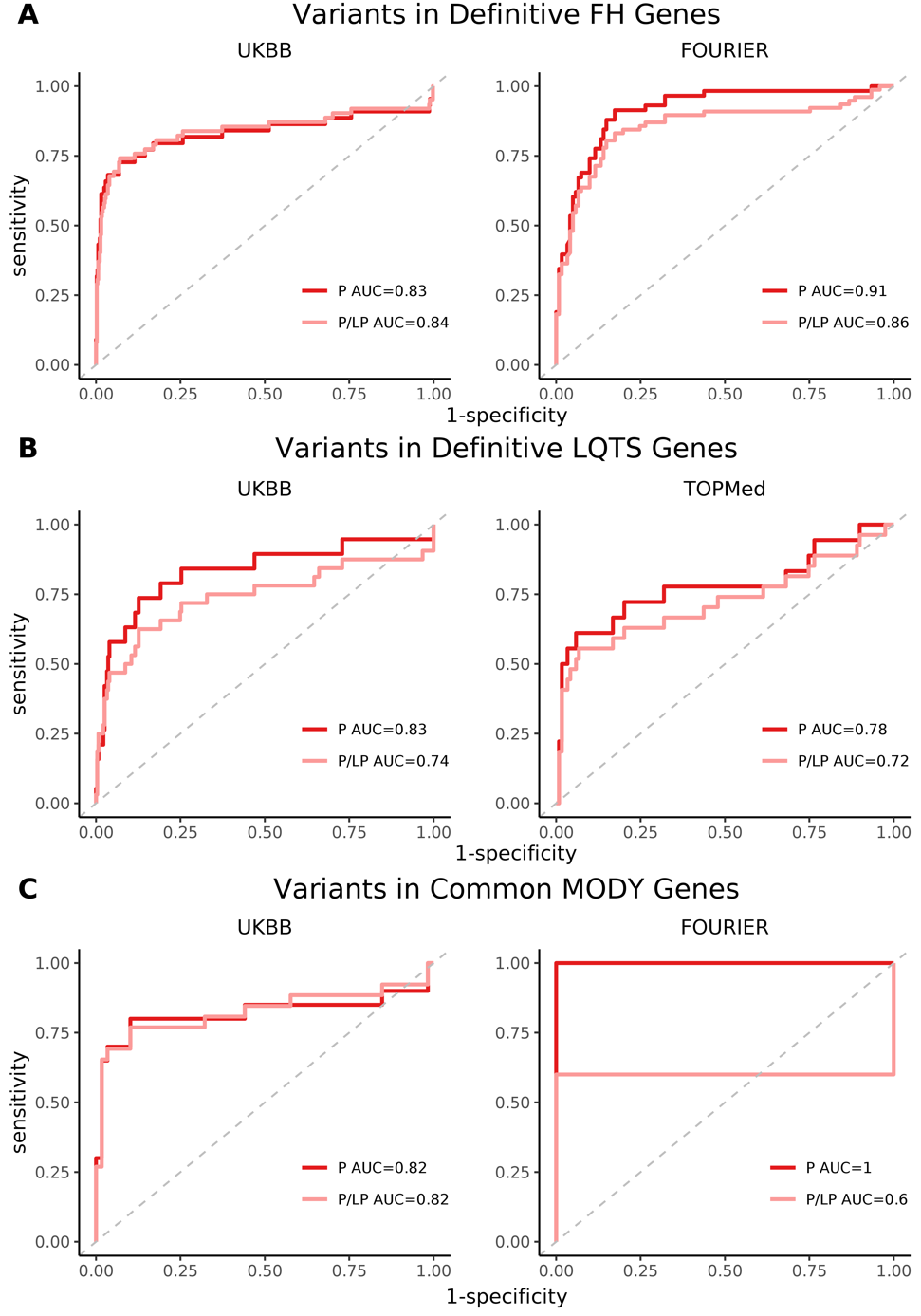

Row A includes variants within familial hypercholesterolemia (FH) genes recommended for secondary findings return (*LDLR, APOB, PCSK9*); Row B includes variants within the long-QT syndrome (LQTS) genes recommended for secondary findings return (*KCNQ1, KCNH2, SCN5A)*; Row C includes variants within common maturity-onset diabetes of the young (MODY) genes (*HNF1A, HNF1B, HNF4A, GCK*). Column 1 includes variants from the UK Biobank, the primary cohort; Column 2 includes variants from FOURIER and TOPMed, the replication cohorts, with removal of variants found in the UK Biobank. ROC curves represent the ability of effect size in discriminating pathogenic variants as defined by ClinVar. The red curve includes “Pathogenic” variants only; the pink curve includes “Pathogenic” and “Likely pathogenic” variants combined; AUC values are reported in the legend.

**Supplemental Acknowledgements**

**UK Biobank**: Use of UK Biobank data was performed under application number 17488 and was

approved by the local Massachusetts General Hospital institutional review board.

**Trans-Omics in Precision Medicine (TOPMed) program**: Whole genome sequencing for the Trans-Omics in Precision Medicine (TOPMed) program was supported by the National Heart, Lung and Blood Institute (NHLBI). Whole genome sequencing for “NHLBI TOPMed: the Atherosclerosis Risk in Communities” (phs001211.v1.p1) was performed at the Broad Institute of MIT and Harvard (3R01HL092577-06S1) and Baylor Human Genome Sequencing Center (3U54HG003273-12S2, HHSN268201500015C). Whole genome sequencing for “NHLBI TOPMed: Genetics of Cardiometabolic Health in the Amish” (phs000956.v1.p1) was performed at the Broad Institute of MIT and Harvard (3R01HL121007-01S1). Whole genome sequencing for “NHLBI TOPMed: Mount Sinai BioMe Biobank” (phs001644.v1.p1) was performed at the Baylor Human Genome Sequencing Center (HHSN268201600033I) and McDonnell genome institute (HHSN268201600037I). Whole genome sequencing for “NHLBI TOPMed: Cleveland Family Study - Whole genome sequencing Collaboration” (phs000954.v1.p1) was performed at the University of Washington northwest genomics center (3R01HL098433-05S1, HHSN268201600032I). Whole genome sequencing for “NHLBI TOPMed: Cardiovascular Health Study” (phs001368.v1.p1) was performed at the Baylor Human Genome Sequencing Center (3U54HG003273-12S2, HHSN268201500015C, HHSN268201600033I). Whole genome sequencing for “NHLBI TOPMed: Framingham Heart Study” (phs000974.v1.p1) was performed at the Broad institute of MIT and Harvard (3R01HL092577-06S1, 3U54HG003067-12S2). Whole genome sequencing for “NHLBI TOPMed: The Jackson Heart Study” (phs000964.v1.p1) was performed at the University of Washington Northwest Genomics Center (HHSN268201100037C). Whole genome sequencing for “NHLBI TOPMed: Multi-Ethnic Study of Atherosclerosis” (phs001416.v1.p1) was performed at the Broad Institute of MIT and Harvard (3U54HG003067-13S1). Whole genome sequencing for “NHLBI TOPMed: Womens Health Initiative” (phs001237.v1.p1) was performed at the Broad Institute of MIT and Harvard (HHSN268201500014C). Core support including read mapping and genotype calling, along with variant quality metrics and filtering were provided by the TOPMed Informatics Research Center (3R01HL-117626-02S1; contract HHSN268201800002I). Phenotype harmonization, data management, sample-identity QC, and general study coordination were provided by the TOPMed Data Coordinating Center (3R01HL-120393-02S1; contract HHSN268201800001I). We gratefully acknowledge the studies and participants who provided biological samples and data for TOPMed.

**TOPMed Study Specific Acknowledgements**:

*Amish: Genetics of Cardiometabolic Health in the Amish*

The TOPMed component of the Amish Research Program was supported by NIH grants R01

HL121007, U01 HL072515, and R01 AG18728. See publication: PMID: 18440328

*BioMe: BioMe Biobank at Mount Sinai*

The Mount Sinai BioMe Biobank has been supported by The Andrea and Charles Bronfman

Philanthropies and in part by Federal funds from the NHLBI and NHGRI (U01HG00638001;

U01HG007417; X01HL134588). We thank all participants in the Mount Sinai Biobank. We also

thank all our recruiters who have assisted and continue to assist in data collection and

management and are grateful for the computational resources and staff expertise provided by

Scientific Computing at the Icahn School of Medicine at Mount Sinai.

ARIC: *Atherosclerosis Risk in Communities study*

The Atherosclerosis Risk in Communities study has been funded in whole or in part with Federal

funds from the National Heart, Lung, and Blood Institute, National Institutes of Health,

Department of Health and Human Services (contract numbers HHSN268201700001I,

HHSN268201700002I, HHSN268201700003I, HHSN268201700004I and

HHSN268201700005I). The authors thank the staff and participants of the ARIC study for their

important contributions.

WGS for “NHLBI TOPMed: Atherosclerosis Risk in Communities (ARIC) Study” (phs001211) was performed at the Baylor College of Medicine Human Genome Sequencing Center (HHSN268201500015C and 3U54HG003273-12S2) and the Broad Institute for MIT and Harvard (3R01HL092577- 06S1). Centralized read mapping and genotype calling, along with variant quality metrics and filtering were provided by the TOPMed Informatics Research Center (3R01HL-117626-02S1). Phenotype harmonization, data management, sample-identity QC, and general study coordination, were provided by the TOPMed Data Coordinating Center (3R01HL- 120393- 02S1). We gratefully acknowledge the studies and participants who provided biological samples and data for TOPMed.

*CHS: Cardiovascular Health Study*

Cardiovascular Health Study: This research was supported by contracts HHSN268201200036C,

HHSN268200800007C, HHSN268201800001C, N01HC55222, N01HC85079, N01HC85080,

N01HC85081, N01HC85082, N01HC85083, N01HC85086, 75N92021D00006 and grants U01HL080295 and U01HL130114 from the National Heart, Lung, and Blood Institute (NHLBI), with additional contribution from the National Institute of Neurological Disorders and Stroke (NINDS). Additional support was provided by R01AG023629 from the National Institute on Aging (NIA). A full list of principal CHS investigators and institutions can be found at CHS-NHLBI.org. The content is solely the responsibility of the authors and does not necessarily represent the official views of the National Institutes of Health.

*CFS: Cleveland Family Study*

The Cleveland Family Study has been supported in part by National Institutes of Health grants

[R01-HL046380, KL2-RR024990, R35-HL135818, and R01-HL113338].

*FHS: Framingham Heart Study*

The Framingham Heart Study (FHS) acknowledges the support of contracts NO1-HC-25195, HHSN268201500001I and 75N92019D00031 from the National Heart, Lung and Blood Institute and grant supplement R01 HL092577-06S1 for this research. We also acknowledge the dedication of the FHS study participants without whom this research would not be possible. Dr. Vasan is supported in part by the Evans Medical Foundation and the Jay and Louis Coffman Endowment from the Department of Medicine, Boston University School of Medicine.

*JHS: Jackson Heart Study*

The Jackson Heart Study (JHS) is supported and conducted in collaboration with Jackson State

University (HHSN268201800013I), Tougaloo College (HHSN268201800014I), the Mississippi

State Department of Health (HHSN268201800015I) and the University of Mississippi Medical

Center (HHSN268201800010I, HHSN268201800011I and HHSN268201800012I) contracts

from the National Heart, Lung, and Blood Institute (NHLBI) and the National Institute on Minority Health and Health Disparities (NIMHD). The authors also wish to thank the staffs and

participants of the JHS.

The views expressed in this manuscript are those of the authors and do not necessarily represent the views of the National Heart, Lung, and Blood Institute; the National Institutes of Health; or the U.S. Department of Health and Human Services.

*MESA: Multi-Ethnic Study of Atherosclerosis*

MESA and the MESA SHARe project are conducted and supported by the National Heart, Lung, and Blood Institute (NHLBI) in collaboration with MESA investigators. Support for MESA is provided by contracts 75N92020D00001, HHSN268201500003I, N01-HC-95159, 75N92020D00005, N01-HC-95160, 75N92020D00002, N01-HC-95161, 75N92020D00003, N01-HC-95162, 75N92020D00006, N01-HC-95163, 75N92020D00004, N01-HC-95164, 75N92020D00007, N01-HC-95165, N01-HC-95166, N01-HC-95167, N01-HC-95168, N01-HC-95169, UL1-TR-000040, UL1-TR-001079, UL1-TR-001420. Funding for SHARe genotyping was provided by NHLBI Contract N02-HL-64278.  Genotyping was performed at Affymetrix (Santa Clara, California, USA) and the Broad Institute of Harvard and MIT (Boston, Massachusetts, USA) using the Affymetrix Genome-Wide Human SNP Array 6.0. MESA Family is conducted and supported by the National Heart, Lung, and Blood Institute (NHLBI) in collaboration with MESA investigators. Support is provided by grants and contracts R01HL071051, R01HL071205, R01HL071250, R01HL071251, R01HL071258, R01HL071259, by the National Center for Research Resources, Grant UL1RR033176. The provision of genotyping data was supported in part by the National Center for Advancing Translational Sciences, CTSI grant UL1TR001881, and the National Institute of Diabetes and Digestive and Kidney Disease Diabetes Research Center (DRC) grant DK063491 to the Southern California Diabetes Endocrinology Research Center.

*WHI: Women’s Health Initiative*

The WHI program is funded by the National Heart, Lung, and Blood Institute, National Institutes of Health, U.S. Department of Health and Human Services through contracts 75N92021D00001, 75N92021D00002, 75N92021D00003, 75N92021D00004, 75N92021D00005.

**TOPMed Omics Support Acknowledgements**

| **TOPMed Accession Number** | **TOPMed Project** | **Parent Study** | **TOPMed Phase** | **Omics Center** | **Omics Support** | **Omics Type** |
| --- | --- | --- | --- | --- | --- | --- |
| phs000956 | Amish | Amish | 1 | Broad Genomics | 3R01HL121007-01S1 | WGS |
| phs001211 | AFGen | ARIC AFGen | 1 | Broad Genomics | 3R01HL092577-06S1 | WGS |
| phs001211 | VTE | ARIC | 2 | Baylor | 3U54HG003273-12S2 / HHSN268201500015C | WGS |
| phs001644 | AFGen | BioMe AFGen | 2.4 | MGI | 3UM1HG008853-01S2 | WGS |
| phs001644 | BioMe | BioMe | 3 | Baylor | HHSN268201600033I | WGS |
| phs001644 | BioMe | BioMe | 3 | MGI | HHSN268201600037I | WGS |
| phs000954 | CFS | CFS | 1 | NWGC | 3R01HL098433-05S1 | WGS |
| phs000954 | CFS | CFS | 3.5 | NWGC | HHSN268201600032I | WGS |
| phs001368 | CHS | CHS | 3 | Baylor | HHSN268201600033I | WGS |
| phs001368 | VTE | CHS VTE | 2 | Baylor | 3U54HG003273-12S2 / HHSN268201500015C | WGS |
| phs000974 | AFGen | FHS AFGen | 1 | Broad Genomics | 3R01HL092577-06S1 | WGS |
| phs000974 | FHS | FHS | 1 | Broad Genomics | 3U54HG003067-12S2 | WGS |
| phs000964 | JHS | JHS | 1 | NWGC | HHSN268201100037C | WGS |
| phs001416 | AA_CAC | MESA AA_CAC | 2 | Broad Genomics | HHSN268201500014C | WGS |
| phs001416 | MESA | MESA | 2 | Broad Genomics | 3U54HG003067-13S1 | WGS |
| phs001237 | WHI | WHI | 2 | Broad Genomics | HHSN268201500014C | WGS |
| ARIC: Atherosclerosis Risk in Communities (ARIC) study, Amish: Genetics of Cardiometabolic Health in the Amish, Baylor: Baylor College of Medicine Human Genome Sequencing Center, BioMe: Mount Sinai BioMe Biobank, Broad Genomics: Broad Institute Genomics Platform, CFS: Cleveland Family Study, CHS: Cardiovascular Health Study, FHS: Framingham Heart Study, JHS: Jackson Heart Study, MESA: Multi-Ethnic Study of Atherosclerosis, MGI: McDonnell Genome Institute, NWGC: Northwest Genomics Center, TOPMed: Transomics for Precision Medicine, WGS: Whole Genome Sequencing, WHI: Women’s Health Initiative | | | | | | |

3.	UKB : Data-Field 42001. https://biobank.ndph.ox.ac.uk/showcase/field.cgi?id=42001.
